# The required size of cluster randomized trials of non-pharmaceutical interventions in epidemic settings

**DOI:** 10.1101/2021.07.12.21260375

**Authors:** Justin K. Sheen, Johannes Haushofer, C. Jessica E. Metcalf, Lee Kennedy-Shaffer

**Affiliations:** Department of Ecology and Evolutionary Biology, Princeton University, Princeton, NJ, USA; Department of Economics, Stockholm University, Stockholm, Sweden; Research Institute of Industrial Economics, Stockholm, Sweden; Max Planck Institute for Collective Goods, Bonn, Germany; Jain Family Institute, New York, USA; School of Public and International Affairs, Princeton University, Princeton, NJ, USA; Department of Mathematics and Statistics, Vassar College, Poughkeepsie, NY, USA

**Author notes:** Correspondence: Lee Kennedy-Shaffer.

**Keywords:** non-pharmaceutical interventions, power, reproduction number, sample size, SARS-CoV-2

## Abstract

To control the SARS-CoV-2 pandemic and future pathogen outbreaks requires an understanding of which non-pharmaceutical interventions are effective at reducing transmission. Observational studies, however, are subject to biases, even when there is no true effect. Cluster randomized trials provide a means to conduct valid hypothesis tests of the effect of interventions on community transmission. While they may only require a short duration, they often require large sample sizes to achieve adequate power. However, the sample sizes required for such tests in an outbreak setting are largely undeveloped and the question of whether these designs are practical remains unanswered. We develop approximate sample size formulae and simulation-based sample size methods for cluster randomized trials in infectious disease outbreaks. We highlight key relationships between characteristics of transmission and the enrolled communities and the required sample sizes, describe settings where cluster randomized trials powered to detect a meaningful true effect size may be feasible, and provide recommendations for investigators in planning such trials. The approximate formulae and simulation banks may be used by investigators to quickly assess the feasibility of a trial, and then more detailed methods may be used to more precisely size the trial. For example, we show that community-scale trials requiring 220 clusters with 100 tested individuals per cluster are powered to identify interventions that reduce transmission by 40% in one generation interval, using parameters identified for SARS-CoV-2 transmission. For more modest treatment effects, or settings with extreme overdispersion of transmission, however, much larger sample sizes are required.

## 1 Introduction

Since its emergence in late 2019, the pandemic SARS-CoV-2 virus has spread globally, resulting in millions of deaths.^1^ Before vaccines were developed and authorized, policy responses to control the spread of the virus relied on non-pharmaceutical interventions (NPIs). NPIs—including, among other measures, mask mandates, school and business closures, and restrictions on travel—have the potential to reduce transmission of the virus. However, which specific NPIs have an effect on transmission remains largely uncertain, while their economic or psychological costs may be substantial. This presents a challenge for policy design. Retrospective statistical analyses of reported cases or deaths,^2–4^ in some cases supplemented with mobility data,^5,6^have provided one means to estimate these impacts, but the conclusions are far from clear. In the first waves of the pandemic, school closures, for example, were found to have relatively minor effects in various types of studies.^2,3,7^ However, more recent analyses suggested that school closure might reduce transmission, perhaps by as much as 15%.^4, 8–11^

Ultimately, while this body of work has helped inform the policy landscape, important gaps remain. Different interventions often overlap in timing, and communities that adopt particular interventions may be similar in other ways, resulting in confounding that biases estimates, even under the null hypothesis of no intervention effect, and challenges the validity of hypothesis tests.^12^ In addition, since these observational studies generally rely on the number of observations available in existing data sets, their power to detect meaningful effect sizes may be low or uncertain.

RCTs are widely used to evaluate the impact of interventions on infectious diseases, as seen in recent vaccine trials with tens of thousands of participants.^13,14^ While the first trials for SARS-CoV-2 vaccines focused on estimating direct effects on individual-level protection, cluster randomized trials (cRCTs) can also provide valuable insight into indirect and total effects of a vaccination regimen in a community.^15,16^ For NPIs, clusters are the natural scale of analysis, as many interventions are implemented at this scale—e.g., by school districts or municipalities—and as both direct and indirect protection are of interest in policy design.

Random allocation of interventions to different communities with short lags has been proposed as an approach to assess whether interventions affect transmission.^17,18^ Indeed, because transmission is rapid, effects can be evaluated in a matter of weeks. Due to uncertainty in results and the short trial duration (and thus short delay before all communities can receive the intervention), equipoise and community acceptability might be relatively high for such trials in an epidemic setting. Despite these advantages, these RCTs might still incur substantial costs and require significant coordination, implementation, and testing. These logistical challenges grow as the number of intervention units increases. Thus, the degree to which deploying RCTs to evaluate NPIs is a useful policy tool will depend on the sample size required to detect, with adequate power, a meaningful reduction in transmission.

Simulation approaches have been previously used to evaluate the statistical power of cRCTs evaluating vaccination at both individual and cluster scales.^19–22^ This allows investigators to size cRCTs to have a desired power to detect a specified true effect size of interest (i.e., reduction in transmission). Although estimators have different properties and interpretations depending on the phase of the epidemic—e.g., by capturing more indirect effects or having less impact when there is more pre-existing immunity— this allows hypothesis tests to be appropriately powered.^21,22^ Here, we build on these results to provide estimates for the number of clusters and number of individuals measured within each cluster needed to test the effectiveness of an NPI, with the aim of bounding the feasibility of such analyses.

We provide investigators with several tools to estimate the required sample size of a cRCT powered to test the effect of an NPI on epidemic transmission. These include approximate formulae that can be used to size the trial, and simulation results that inform how power depends on key parameters. For more precise sample size and power calculations, simulations adapted to the context of the trial under consideration will be necessary. The results presented here, however, can provide a baseline for assessing whether a trial may be feasible with a reasonable sample size and can provide a starting point for more specific investigations.

## 2 Methods

### 2.1 Trial Design

We consider cRCTs where there are *N* = *N*_1_ + *N*_0_ total clusters enrolled, with *N*_1_ in the intervention arm and *N*_0_ in the control arm. At a specified day *t* after the introduction of infections into the clusters, one round of sampling is performed, sampling *m*_0_ individuals from each cluster and testing them for the infection. In the simulations, we find the time *t* such that the average proportion of infectious individuals is as desired, averaged across clusters. We then set this value *t* for use in all clusters. At that point, the intervention begins in the intervention arm clusters, a randomly-selected subset of half of the study clusters. A set amount of time later (in our main results, equal to one generation interval of the infection, but a range of times are explored in Section 3.2.2), another round of sampling occurs, this time sampling *m*_1_ individuals from each cluster and testing them. We denote this time point by *t* + 1. We assume that the test accurately identifies infectious individuals. More complex models may be used to adjust for known imperfect sensitivity and specificity, as has been done in other settings.^12,23^

In some cases where clusters are relatively small, it may be reasonable to assume that everyone or nearly everyone in the cluster will have their outcome measured. For example, some schools, universities, long-term care facilities, and workplaces have proposed or implemented universal testing strategies.^24–27^ We consider this setting first, followed by settings where a simple random sample (without replacement) is chosen for testing, independently at each time point.

Each cluster *j* then has two values associated with it: *Y*_*j,t*_, the number of sampled individuals who test positive in the first round of sampling (pre-intervention); and *Y*_*j,t*+1_, the number of sampled individuals who test positive in the second round of sampling (post-intervention). We conduct analysis using test statistics based on the quantity 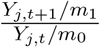 from each cluster, e.g., by comparing the mean of this statistic in the intervention arm to that in the control arm. This statistic estimates the reproductive number, *R*_*t*_, in the cluster, so the difference in means estimates the reduction in the transmission rate.^17^

This statistic may not always be unbiased, due to adjustments made for zero-case clusters, asymmetric effects of the progress of the epidemic prior to time *t*, and the continuous-time nature of transmissions. However, in a randomized trial, under the null hypothesis of no effect of intervention, the statistic has zero expectation. Thus, hypothesis tests based on this statistic are valid. In the sample size calculations to follow, we size the trial based on the true effect of intervention on the reproductive number. While the power and sample size calculations presented here are based on hypothesis tests using this statistic, they may be reasonable approximations for other test statistics using the same information. While Type I Error is preserved through internal validity, other statistics may have greater power, especially if they avoid any bias in the estimator. In addition, other sampling schemes are possible, including sampling only at time *t* + 1 or additionally using serologic sampling to estimate the number of susceptible individuals at either or both time points. We focus on the setting using only virologic testing at the two time points for power calculations as it may be broadly feasible to implement.

### 2.2 Epidemic Spread Assumptions

Both the development of approximate sample size formulae and the simulations that follow depend on certain assumptions about the epidemic process. First of all, we assume that clusters are independent; that is, there is no transmission between clusters. This may be reasonable if clusters are sufficiently geographically distinct.

Secondly, we assume that once a cluster has its initial infections, the pathogen spreads according to a standard Susceptible-Exposed-Infectious-Recovered (SEIR) model. In the approximations, we use a discrete-time SEIR model with a fixed generation interval. We assume that at time *t*, the proportion of individuals who are infectious is *I*_*t*_. This model assumes a time-varying reproduction number, *R*_*t*_, that proceeds in each community from a common basic reproduction number *R*_0_, changing as the number of susceptible individuals changes, with overdispersion parameterized by *k*. This parameter allows for the epidemic spread to encompass settings ranging from very little variation in transmission across individuals (*k ≥* 1) to a small number of infected individuals being responsible for the vast majority of onward transmission (*k* < 0.1), sometimes referred to as “superspreading”.^28, 29^ The number of individuals in the community is assumed to be sufficiently large compared to the number of infectious individuals at time *t* such that no individual is infected by two infectious individuals in the same generation.

In the simulations, we use a continuous-time stochastic SEIR model. We assume that each exposed individual’s incubation period is drawn from an exponential distribution with a mean of 5.51 days.^30^ We match the simulations done elsewhere,^21,22^ assuming the mean infectious period across individuals is 5 days (from an exponential rather than a gamma distribution). We assume an approximate negative binomial degree distribution for the network structure of each cluster with a mean of 15 contacts per individual and overdispersion parameter *k*. The contact structure does not exactly follow the negative binomial degree distribution because we delete self-loops after using a configuration model (CM) algorithm to create the contact structure.^31^ We also assume a fixed initial number of infections to seed the epidemic according to cluster size (see Table 1). Note that because the epidemics progress stochastically within each cluster, the *R*_*t*_ and *I*_*t*_ values vary between clusters.

**Table 1:** Parameters Used in Simulations

| Parameter | Values |  |  |
| --- | --- | --- | --- |
| $R_0$ | 1.5, 2.0 | | |
| Overdispersion Parameter ($k$) | 0.1, 0.4, 0.7 | | |
| Effect Size (Reduction in Transmission) ($\Delta$) | 20%, 40% | | |
| Cluster Population ($n$) | 100 | 1,000 | 10,000 |
| Number Sampled Per Cluster ($m$) | 10, 50, 100 | 100, 1,000 | 100, 1,000 |
| Mean Infection Prevalence at $t$ ($E[I_t]$) | 2% | 0.5% | 0.5%* |
| Initial Infection Prevalence ($I_0$) | 1% | 0.4% | 0.4% |

\*0.45% for $n = 10,000$ when $k = 0.1$

In the approximations, the intervention can affect the reproduction number and the overdispersion parameter in the clusters where it is implemented. In the simulations, the intervention affects the transmission rate, but not the contact structure in the cluster and thus not the overdispersion of contacts.

### 2.3 Approximate Sample Size Formulae

We consider analysis based on the test statistic comparing the means of 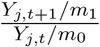 (the ratio of the proportion infected post-intervention to pre-intervention, which approximates the reproduction number when measured one generation interval apart) in the intervention vs. control arms. We present two results based on approximate distributions of the test statistic: (i) an approximate sample size required (in terms of the number of clusters per arm) when there is full testing within each cluster at times *t* and *t* + 1; and (ii) an approximate sample size required when there is sampled testing within each cluster at those times. The former is useful for small cluster sizes, where full or near-full testing is feasible. The latter assumes that the number tested is small compared to the total cluster population, although it focuses on the variability due to post-intervention sampling and ignores the variability due to sampling at time *t*. The latter is thus likely to underestimate the required sample size in many settings, except where the proportion sampled is non-negligible (i.e., greater than 10%). The approximations do not account for the number susceptible, so both methods are most accurate early in an outbreak.

Both of these results are based on a Welch’s two-sample *t*-test for the comparison of two means, with unequal variances. For this test, the required sample size (number of clusters) in each arm, *N*_*i*_, to detect a difference in means of Δ with power 1 − *β* at two-sided significance level *α* solves:^32^

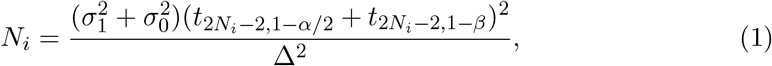

where 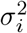 is the variance of the observations in intervention arm *i*, for *i ∈ {*0, 1*}*, and *t*_*DF,ϕ*_ is the *ϕ*-quantile of the *t* distribution with *DF* degrees of freedom. We present variance estimates that can be used in this calculation, with proofs and a full statement of the assumptions and approximations made in the derivation given in Appendix 1.

If the full cluster populations are tested at each time point, then an approximate sample size can be calculated for the difference in the means of 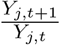 between the intervention arms, where *Y*_*j,t*_ is the number of individuals who test positive at time *t* in cluster *j*. The *t*-test can then be used, with effect size 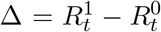 estimated by the difference in means, and variances approximated for each arm *i* by:

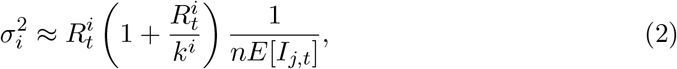

where 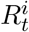 is the time-varying reproduction number at time *t* in intervention arm *i, k*^*i*^ is the overdispersion parameter of transmission in intervention arm *i, n* is the population in each cluster, and *E*[*I*_*j,t*_] is the mean (across clusters) proportion of individuals who are infectious at time *t*.

If the variance of the proportion of individuals who are infectious at time *t* across clusters, *V ar*[*I*_*j,t*_], can be estimated as well, then the calculation should use variances approximated by:

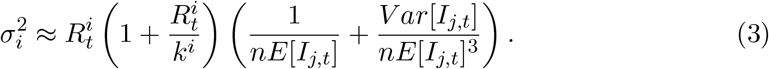

When a small proportion of the population of each cluster is tested at each time point instead (specifically, *m*_0_ individuals per cluster at time *t* and *m*_1_ individuals per cluster at time *t*+1), the effect size 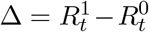 can be approximated by the difference in the means of 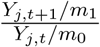. The variances can be approximated by:

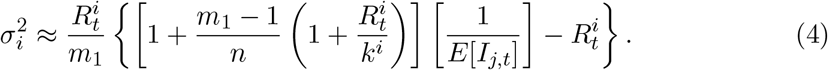

Again, if we can estimate the variance of the proportion of individuals who are infectious at time *t* by *V ar*[*I*_*j,t*_], then the variances can be approximated by:

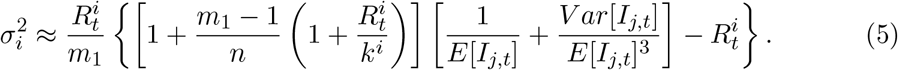

R functions to calculate these values are available at http://www.github.com/jsheen/NPI.

### 2.4 Simulation Setup

For each parameter combination described in Table 1, we first create a simulation bank of cluster simulations with and without an enacted NPI intervention. The simulated epidemic is described above. To approximately align clusters on epidemic time, time *t* is defined as the first day when the mean proportion of infectious people across clusters is approximately equal to the target *E*[*I*_*t*_]. In an epidemic, it would be reasonable to test interventions at similar epidemic time points, but the remaining variability in the number infected allows us to explore the impact of this variance on power. After *t* is identified, the simulation bank is created by interrupting the simulation for each cluster at time *t*. The per-contact daily transmission rate, *β*, is set empirically to give the desired *R*_0_ at the beginning of the simulation. At time *t*, we continue the cluster simulation both with and without an enacted NPI intervention for one generation interval (11 days) and record the number of infectious individuals at this point (denoted time *t* + 1). The generation interval is equal to the ceiling of the sum of the average incubation period and average infectious period.^33,34^ At times *t* and *t* + 1, *m* individuals are sampled and tested within each cluster. We create 3,000 simulations for the simulation bank. Only simulated clusters with at least one infectious individual at time *t* are kept, so we implicitly assume there is at least one infectious individual in each cluster. Example trajectories of the simulations are shown in Figure 1.

**Figure 1:**
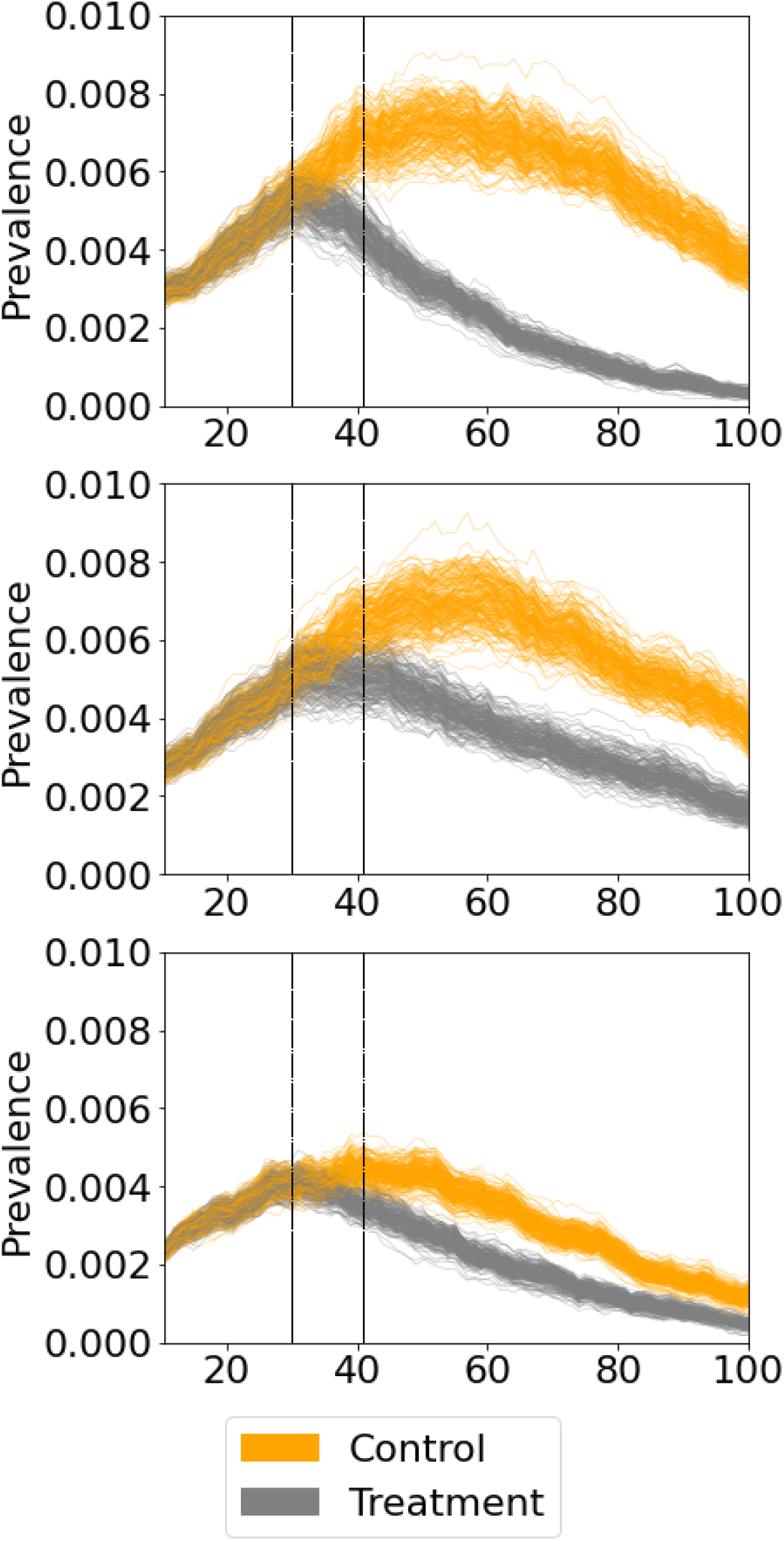
Simulated trajectories of a randomized control trial of a non-pharmaceutical intervention. Each arm has 100 clusters, with average basic reproduction number under control *R*_0_ = 1.5, overdispersion parameter *k* = 0.4 (top and middle panels) or *k* = 0.1 (bottom panel, indicating more overdispersed transmission), and cluster size *n* = 1, 000. On day 30, the intervention begins, which reduces the transmission rate, *β*, by 40% (top panel) or 20% (middle and bottom panels). The dashed lines represent the day of intervention (*t*) and the day of sampling, one generation interval after intervention (*t* + 1).

To find the number of clusters in each arm of the trial needed to achieve approximately 80% power when *α* = 0.05, we use a binary search algorithm with a minimum and maximum number of clusters of 1 and 1,000, respectively. At each iteration of the algorithm, 10,000 trial simulations are performed by choosing a given number of clusters from each of the treatment arm and the control arm in the simulation bank. The empirical power is then calculated after sampling *m* individuals from each cluster and a two-sample Welch’s t-test is performed on the quantity 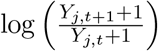. Note that to avoid undefined test statistic values, we add one infected individual at each time point to each cluster. This binary search algorithm implicitly assumes that the relationship between power and number of clusters monotonically increases. Algorithm pseudocode for the creation of the simulation bank and binary search algorithm for *N* can be found in Appendix 2, along with comments on the stability of the algorithm (see Figure S1).

We further extend these simulation results in two ways: (i) we provide *N* when the post-intervention testing time point is either two or three generation intervals after *t* instead of one generation interval; and (ii) we provide simulation results when matching clusters into pairs depending on the number of susceptible individuals at time *t*, as well as the number of non-infectious individuals at time *t*, within each cluster, using a matched-pairs *t*-test. We use a greedy matching algorithm and randomly assign one cluster of each pair to either treatment or control.

We use the *EoN* python package to simulate the epidemic.^35^ Code used for simulations is provided at http://www.github.com/jsheen/NPI.

## 3 Results

We present results from both the approximate sample size calculations and simulations that target an empirical power of 80% to detect a specified effect size with two-sided significance level *α* = 0.05 for the parameter combinations described in Table 1.

### 3.1 Approximate Sample Size Requirements

The required sample size depends on features of the transmission of infection, the sizes of the cluster populations and samples, and the effect size studied. These relationships are illustrated using the approximate variance formulae.

#### 3.1.1 Approximations Under Full Measurement

In cases where the outcome (infection) is measured in everyone in each cluster, we can use equation (2) to estimate the variance. The variances (and thus required sample sizes) increase as the reproduction numbers, 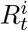, decrease or as the overdispersion parameter, *k*^*i*^, decreases. Note that decreasing *k*^*i*^ corresponds to more overdispersion and thus more variability in the change in the number of infections over one generation. In addition, the variance increases as the average proportion of infected individuals per cluster at time *t* decreases. These relationships are plotted in Figure 2, which shows the number of clusters per arm required for 80% power to detect a reduction in transmission of 40% at significance level *α* = 0.05. Figure 2A shows that, for a given cluster size and expected proportion of infections at enrollment, there is a slight decrease in the required sample size as 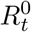 increases and a substantial decrease in the required sample size as *k* increases (less overdispersion). Figure 2B shows that, for fixed 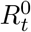 and *k* (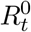 = 1.5 and *k* = 0.4 are shown), the required number of clusters decreases as the expected proportion of the population infected at enrollment, *E*[*I*_*t*_], increases and as the cluster size (and thus number sampled) increases. When the expected number of infections per cluster at time *t* falls below approximately two, the required sample size increases dramatically.

**Figure 2:**
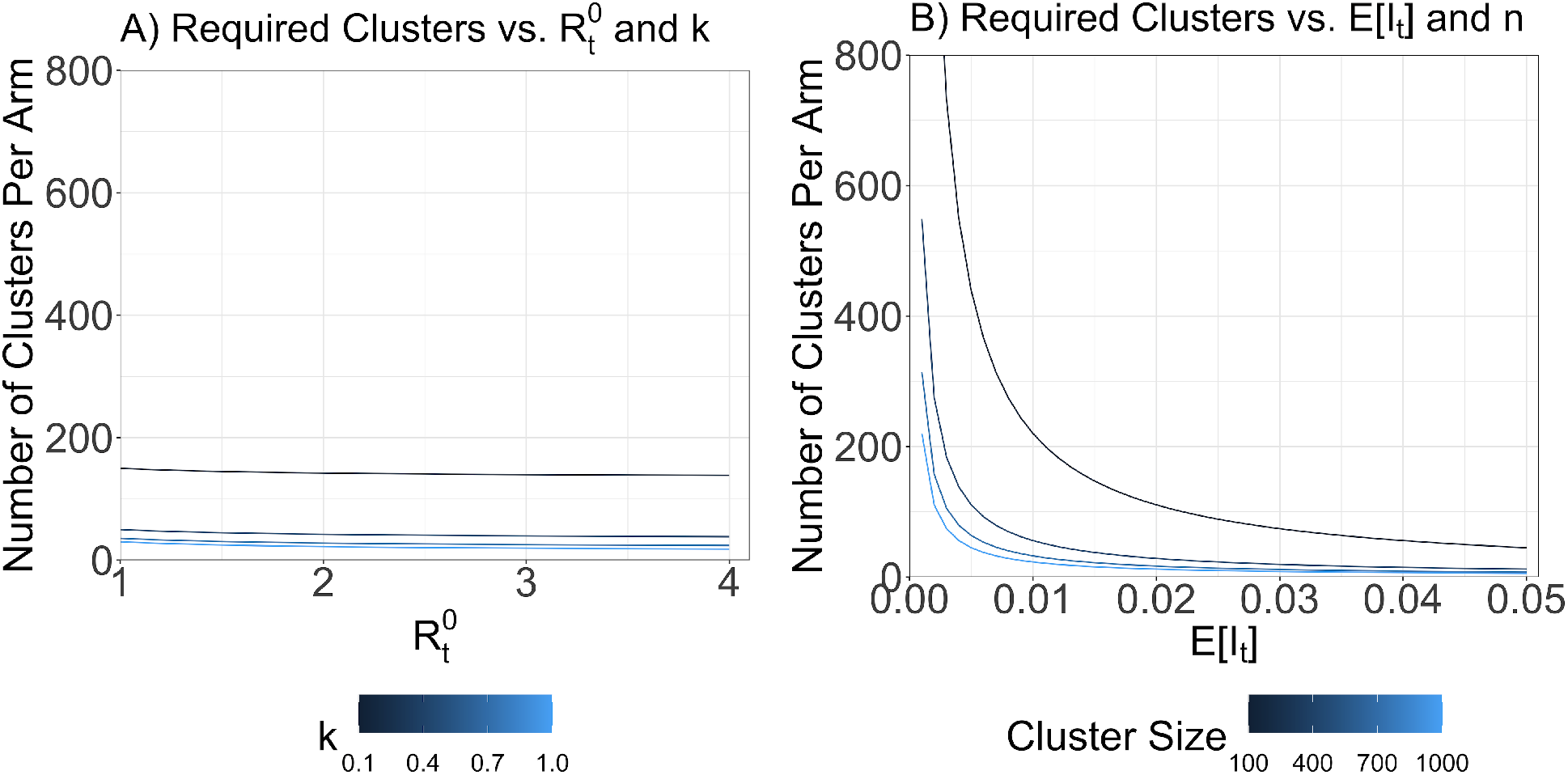
When all individuals are tested in each cluster, the required number of clusters per arm decreases as the reproduction number, overdispersion parameter, proportion infected at time of intervention, or cluster size increase. Approximate number of clusters per arm required for 80% power to detect an effect size of 40% at *α* = 0.05 vs. (**A**) reproduction number under control (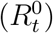) and overdispersion parameter (*k*) for fixed cluster size *n* = 1, 000 and expected proportion of infections at enrolment *E*[*I*_*t*_] = 0.005 and vs. (**B**) *E*[*I*_*t*_] and *n* for fixed 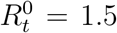 = 1.5 and *k* = 0.4. Note that higher values of *k* correspond to more overdispersed transmission.

#### 3.1.2 Approximations Accounting for Sampling

When sampling within each cluster is accounted for, similar relationships are observed between 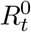, *k*, and the required sample size calculated using equations (1) and (4). Figure 3A shows these relationships for a cluster size of *n* = 10, 000 and sampling *m* = 100 individuals, with *E*[*I*_*t*_] = 0.005, and with an effect size (transmission reduction) of 40%. Because of the larger cluster size, the spread of infections is more deterministic, leading to a smaller effect of overdispersion. Figure 3B shows how the effect size affects the required sample size for fixed *k* = 0.4.

**Figure 3:**
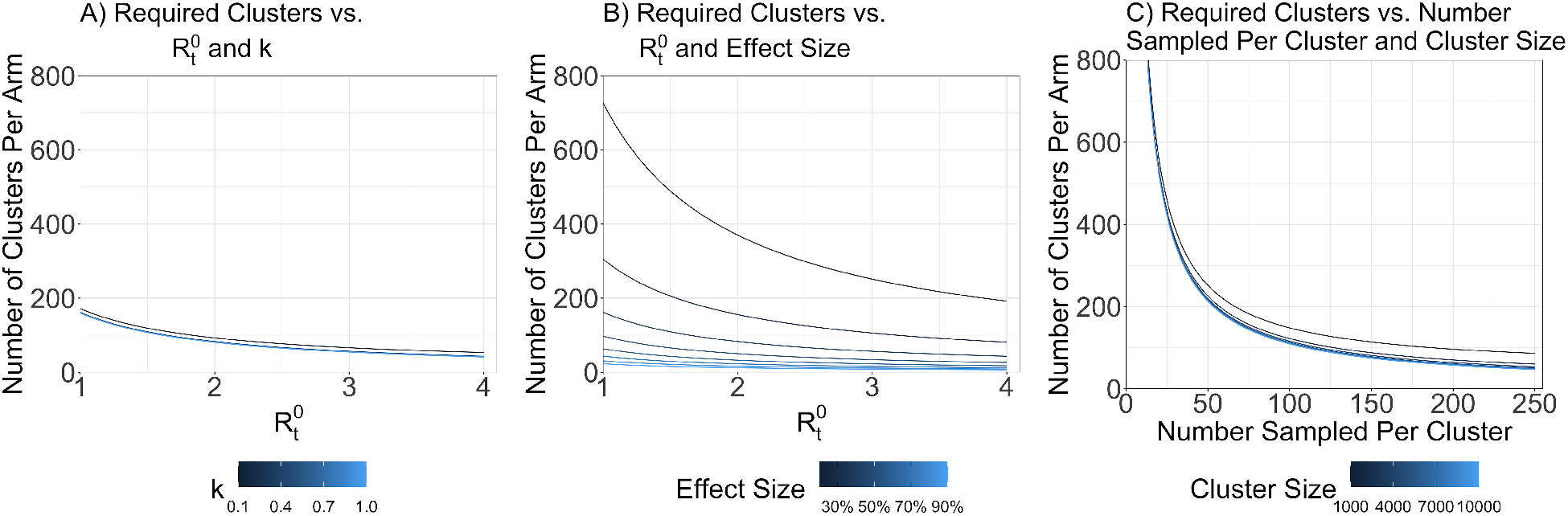
When a sample of individuals in each cluster are tested, the approximate required number of clusters per arm decreases as the reproduction number, overdispersion parameter, effect size, cluster size, or number of individuals sampled per cluster increase. Approximate number of clusters per arm required (as calculated by equations (1) and (4)) for 80% power to detect an effect size of 40% (**A**,**C**) or as specified (**B**) at *α* = 0.05 vs. (**A**) reproduction number under control 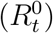 and overdispersion parameter (*k*) for fixed cluster size *n* = 10, 000 and number sampled per cluster *m* = 100; vs. (**B**) 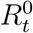 and effect size for fixed *n* = 10, 000, *m* = 100, and *k* = 0.4; and vs. (**C**) *n* and *m* for fixed 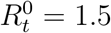 = 1.5, and *k* = 0.4. In all panels, the expected proportion of the population infected at enrollment is *E*[*I*_*t*_] = 0.005.

With this approximation, we can also examine the relationship between the number of individuals sampled per cluster, the cluster size, and the required sample size. Figure 3C illustrates these relationships when 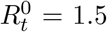 = 1.5, *k* = 0.4, the reduction in transmission due to intervention is 40%, and *E*[*I*_*t*_] = 0.005. Note that these approximations ignore any finite sample corrections. When the number sampled per cluster is a large proportion of the cluster size (i.e., 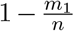 is meaningfully less than 1), this difference is likely to be meaningful.^36^ For reference, the approximate sample size if the full cluster is tested with these parameters ranges from six clusters per arm if *n* = 10, 000 to 45 clusters per arm if *n* = 1, 000, which represent (approximately) the minimum number of clusters for less-than-complete sampling.

There are two key effects of an increase in cluster size, holding all other parameters fixed: (i) stochastic effects in epidemic spread are less pronounced, leading to more similar epidemic trajectories across clusters; and (ii) the number of individuals sampled represents a smaller proportion of the cluster population. The former effect tends to decrease the variance of the test statistic, while the latter effect tends to increase the variance of the test statistic. Thus, it is difficult to describe a general rule governing the relationship between cluster size and required sample size. In the approximations shown in 3C, the latter effect is ignored, so the estimated required number of clusters decreases as cluster size increases.

As the number of individuals sampled per cluster increases, there is a reduction in the required number of clusters per arm. However, this exhibits diminishing returns as it increases, indicating an eventual tradeoff between the number of clusters required per arm and the total number of samples required per arm, as is common in cRCTs.^20^ This figure likely underestimates the value of increased testing per cluster, especially for relatively large fractions sampled, as it ignores finite population corrections.

### 3.2 Sample Size Requirements from Simulations

Estimated required sample sizes to get the desired empirical power are calculated in simulations as well. A full set of results from these simulations are shown in Supplementary Tables S1–S17. Here, we focus on the relationships between the key parameters and the required sample size.

#### 3.2.1 Sample Sizes Sampling One Generation After Intervention

Similar to the approximation results, simulation results demonstrate that, in general, the required sample size decreases as the overdispersion parameter increases, 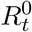 decreases, effect size increases, and the number of individuals sampled per cluster increases (Figure 4).

**Figure 4:**
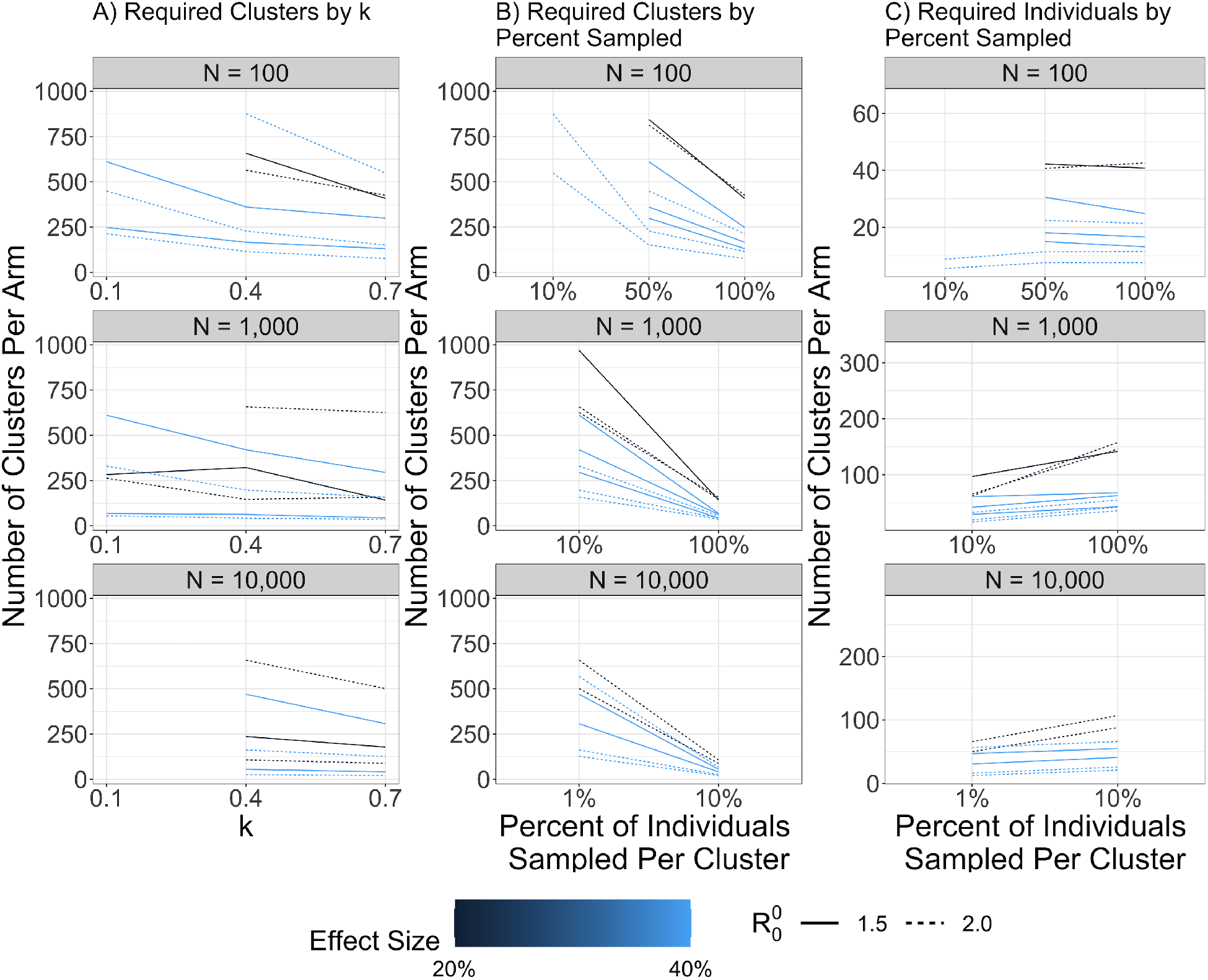
The required number of clusters per arm to achieve a desired empirical power in simulations depends on the overdispersion parameter, reproduction number, effect size, cluster size, and percent of individuals in each cluster who are sampled. Number of clusters per arm required (**A**,**B**) and number of individuals per arm required (**C**) to achieve 80% empirical power with a significance level of *α* = 0.05 in 10000 simulated trials vs. overdispersion parameter *k* (**A**) or percent of individuals sampled per cluster (**B**,**C**). Effect size and basic reproduction number at the start of the outbreak, 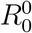, are varied within each panel. In **A**, each percent of individuals sampled has a unique line, leading to larger differences even when effect size is held fixed.

Figure 4A shows that as overdispersion decreases (*k* increases), the required sample size decreases. Moreover, as shown in Tables S1–S3, as *k* increases, 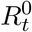 at time of intervention generally increases as well, even for fixed 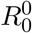 at the start of the simulated outbreak. Because of this, the relationship between *k* and 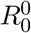 and the required sample size is even more pronounced in the simulation results. Figure 4B illustrates that the required number of clusters will generally decrease as a greater percentage of the cluster is sampled. This indicates that if testing is easy to conduct, the required number of clusters for a trial can be reduced by increasing the sampling within each cluster. Conversely, if the total number of individuals to be sampled in the trial is fixed (i.e., limited number of tests available), and the number sampled from each cluster and number of clusters required are allowed to vary, Figure 4C illustrates that it is more efficient to sample fewer individuals from a greater number of communities than it is to sample more individuals from a smaller number of communities. This relationship is less clear for small clusters (*n* = 100) where nearly full sampling can occur.

For clusters of size *n* = 100 or 1, 000, the day of NPI intervention for some parameter sets occurred less than four weeks after the start of the epidemic (when *I*_*t*_ = 2% and 0.5% respectively)—but interventions may not always be able to be implemented this quickly. To account for longer delays between the start of the epidemic and day of intervention, we further extend our results by reporting the sample sizes when the day of intervention is one month after the first day of infection in Tables S4 and S5.

#### 3.2.2 Sample Sizes With Greater Lags After Intervention

The formulae and results described above all tested the effect of intervention after one generation interval. Discretizing on this time scale will provide an estimate that approximates transmission, *R*_*t*_, although occurrences of secondary infections prior to the full generation interval will bias estimates of transmission upwards, and occurrences of primary infections occurring after the full generation interval will bias estimates downwards.^37^ Furthermore, because the direct effects continue and indirect effects may increase on short time scales, the effect size in cRCTs in epidemics can increase over time.^21,22^ Eventually, however, the exhaustion of susceptible individuals will lead to a reduction in the effect size as incidence rates become more similar between intervention and control clusters. We explore the effects of the time interval used in our simulations by increasing the lag between intervention and evaluation. The approximations are not well-suited to assess these sample sizes as their assumptions become less reasonable over longer time scales.

Figure 5 shows a drastic increase in power—or decrease in the required sample size— if sampling occurs two generation intervals after intervention compared to sampling one generation interval after intervention.

**Figure 5:**
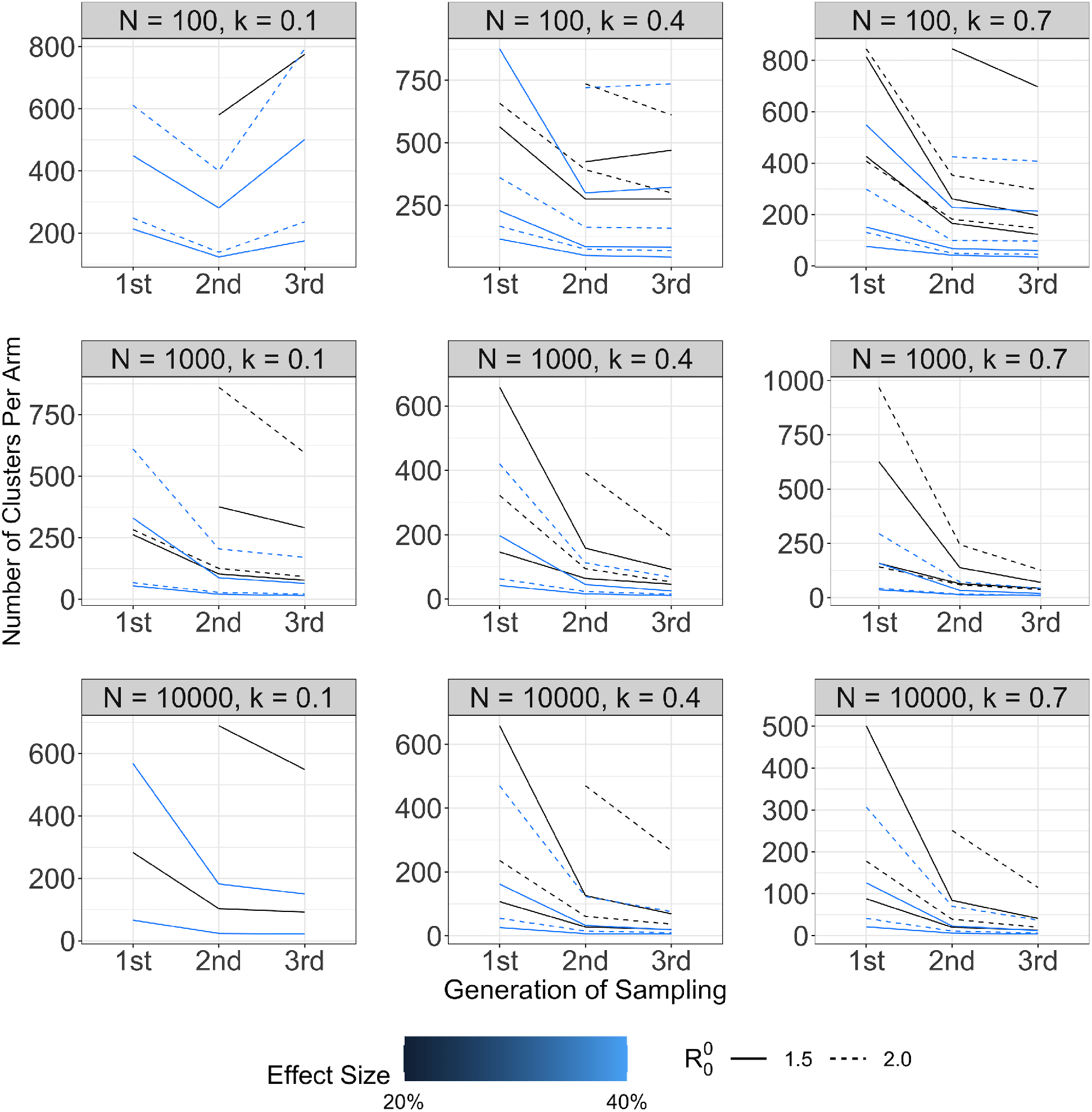
Required sample size generally decreases as time of sampling after intervention increases, except when the number of true infections is low three generation intervals after intervention for smaller cluster sizes. Rows correspond to cluster size, *n* and columns correspond to overdispersion parameter, *k*. Effect size, basic reproduction number at the start of the outbreak, 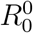, and number sampled, *m*, are varied within each panel. On average, sampling two or three generation intervals after intervention required a sample solely 35% and 31% as large, respectively, as after one generation interval.

However, these results change when we increase the lag further, to sampling three generation intervals after intervention. There is generally a more modest decrease in the required sample size for extending from two to three generation intervals than from one to two generation intervals. Second, for small clusters (*n* = 100), increasing the lag between intervention and day of sampling can even increase the required sample size for certain parameter combinations. As the epidemic progresses, there is eventually a point where the depletion of susceptible individuals leads to a decline in power from increasing the lag, although this time point will depend on the precise combination of parameters. We also find this decrease in power occurs more often when there are fewer people sampled from each cluster. Full results are shown in Tables S6–S11.

#### 3.2.3 Sample Sizes After Matching Clusters

A common approach to increasing power in cRCTs is to match or stratify clusters on baseline covariates to increase balance and reduce variability.^20, 38^ In this case, clusters can be matched on the number of susceptible individuals at the time of intervention (assuming the availability of serological tests) or on the number uninfected at the time of intervention (using the prevalence data collected at time *t*). This matches clusters on *I*_*j,t*_, reducing the effect of the variability in that parameter on the variance of the effect estimate.

We assess the effect of this matching in the simulations, using a matched pairs *t*-test where clusters are matched on the number of susceptible individuals at the time of intervention (see Tables S12–S14. We find evidence of modest benefits from matching when cluster sizes are large. The required sample size generally decreases depending on the parameter combination used; however, on average, the change in required number of clusters for *n* = 1, 000 and *n* = 10, 000 was modest: 5% and 2% reductions in required sample size, respectively.

For clusters of size 100, the required sample size improved for all parameter combinations where we were able to solve for the number of clusters. The average reduction in required number of clusters was 14%. Because of the smaller cluster size, matching on the number of sampled susceptible individuals may be more reliable and thus more informative than matching with larger cluster sizes. More specifically, the number of sampled susceptible individuals in a smaller cluster compared to a larger cluster may give more information about the number of cases in the following generations because it has a larger impact on the transmission dynamics.

We also assess the effect of matching when clusters are matched on the number of non-infectious individuals at the time of intervention (see Tables S15–S17). We find larger benefits from matching on this parameter: the decreases in the required number of clusters for *n* = 100, 1, 000, and 10, 000 were 15%, 12%, and 28% on average, respectively. In general, for each cluster size, the benefits were greater for smaller sample sizes.

When comparing matching clusters on number of susceptible individuals to the number of non-infectious individuals at time of intervention, we find that when *n* = 100 and 1, 000, when not all individuals were tested, matching on the number of noninfectious individuals reduced the sample size compared to matching on the number of susceptible individuals (by 24% and 27% for *n* = 100 and 1000, respectively). When all individuals in each cluster were tested, matching on number of non-infectious individuals increased the sample size compared to matching on susceptible individuals (by 27% and 8% for *n* = 100 and 1000, respectively). When *n* = 10, 000, matching on the number of non-infectious individuals reduced the sample size compared to matching on susceptible individuals by an average of 26%. Thus, using serology testing to capture the number of susceptible individuals at time *t* may be more useful when all participants are tested; when clusters are not fully sampled, in contrast, the number of sampled non-infectious individuals (equivalently, the number of sampled infectious individuals) at time *t* is more useful for matching.

Importantly, we assess the benefits of matching when time of intervention occurs according to a pre-specified *E*[*I*_*t*_] in Table 1. Time of intervention will change the number of individuals of each condition (susceptible, exposed, infectious, and recovered), thus affecting the benefits of matching. The effect of time of intervention on the benefits of matching is not explored.

Gains in power may also be achieved using stratification or adjustment by measured covariates related to the transmission of infections within each community. For cluster-level covariates, analysis would then proceed by regression analysis. The reduction in sample size depends on the correlation between the covariate and the outcome, and we refer readers elsewhere to determine the expected reduction that could be applied to calculated sample sizes.^20, 32^

### 3.3 Comparison of Approximation and Simulation Sample Sizes

The approximation and simulation approaches generate different ways of considering the issue of required sample sizes, with the former illustrating the impact of different contextual features (overdispersion, cluster size) and the latter accounting for more of the variability in the epidemic spread process. Direct comparison between the sample size requirements derived from approximation formulae and from simulations are difficult, primarily because of the progression of the epidemic up to the time of intervention in the simulations. As previously susceptible edges of the contact network have already been infected in some clusters, the distribution of infections in the next generation varies from cluster to cluster. Similar effects may occur with the contact networks, where overdispersion falls over time as the most highly-connected individuals are most likely to have already been infected.^39, 40^

Despite this, the approximations and simulations shown here generally provide required sample sizes that are comparable in magnitude for many settings. When all individuals are tested in each cluster, the approximation performs very well and closely matches the results of simulations. When the variance of infections at time *t* across clusters is ignored, this is likely to result in some underestimation of the variance and thus of the required sample size; the same is true because variation in the 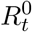 values across clusters is ignored by the approximation.

When only a sample of individuals is tested in each cluster, the approximations diverge further from the simulations. This can occur for a variety of reasons in addition to the parameter mismatch described above: the approximation does not account for sampling prior to the intervention and it does not fully account for the variance in the number of infectious individuals at the time of intervention, *V ar*[*I*_*t*_], and the variance in the actual reproductive number, 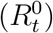, at the time of intervention. Figure S2 shows the wide spread in the number of infectious individuals per cluster, which reflects the overdispersion of the contact structure of the simulated networks. When the outbreak parameters can be well-estimated *a priori*, then, the simulations account for certain heterogeneities that the approximations do not. Because of this, approximations can be used to get an estimate of the feasibility of a trial but should not be the only consideration in powering a trial.

Moreover, due to the stochasticity, some clusters may have zero identified cases at time *t* + 1. This is accounted for in the analysis of the simulations by adding one case to each time point, which may introduce some bias. Additionally, the test statistic inherently is based on a fixed, discrete generation interval, which is not the case in the simulation or in reality. This may lead to the test statistic not estimating the reduction in *R*_*t*_ consistently; however, since the null remains the same in either case, the hypothesis tests remain valid.

## 4 Discussion

To determine whether cRCTs are a practical tool to test the impact of NPIs in epidemic settings, we developed two approximate sample size formulae. We compared these results to simulated outbreaks and developed a simulation bank that can be used to further refine estimates of the required sample size for cRCTs. The simulations can be adapted to specific settings to provide more precise sample size estimation and improve the design of cRCTs.

As an example, we have shown that for settings with communities of 10,000 people, *R*_*t*_ of 1.5 in the absence of intervention, and *k* of 0.4, 80% power to detect a reduction in *R*_*t*_ of 40% due to intervention can be achieved with approximately 220 total clusters (22,000 sampled individuals) in the trial. While this is certainly a large sample size, cRCTs of that order of magnitude have been conducted for large-scale policy interventions,^41^ and individual RCTs with thousands or tens of thousands of participants have occurred to evaluate NPIs and vaccines during this pandemic.^13,14,42, 43^ In particular, if large-scale random testing of individuals is occurring that can be incorporated into the study, sampling large numbers of individuals per cluster may be feasible. As *R*_*t*_ increases, overdispersion decreases, or the effect size increases, this sample size can be reduced while maintaining power.

For communities of 100 people who are all tested (e.g. a workplace), with the same transmission parameters, 80% power to detect a reduction in *R*_*t*_ of 40% due to intervention can also be achieved with approximately 220 total clusters (22,000 sampled individuals) in the trial. If the overdispersion were more extreme, the required sample size would increase drastically; for example, for *k* = 0.1, these trials would require approximately 720 clusters, or 72,000 sampled individuals. If, in addition, 80% power to detect a reduction in *R*_*t*_ of 20% is desired, the sample sizes increases dramatically, requiring approximately 3,500 clusters or 350,000 sampled individuals.

These results use a simple estimator based on virologic testing at two time points, one before and one after the intervention.^17^ More work is needed to determine the properties of this estimator, especially in cases where the epidemic fades out in certain clusters and as the lag between the two testing times changes, as both may bias estimates away from the true transmission reduction. Other estimators may have more desirable properties in estimating specific estimands of interest or in precision. We focus here only on the power of hypothesis tests, the ability to reject the null of no intervention effect for interventions that reduce the reproduction number by a specific amount.

The approximation formulae are limited by the fact that they ignore the variability in the number of active infections at time *t*, and the method that accounts for sampling ignores finite population corrections and sampling variability at time *t*. In addition, these methods ignore the variability in previous infections and the effect those have on future spread on the network, which may serve to overstate the variability in dispersion, especially for small cluster sizes or late time points in the epidemic.^39, 40^ The simulations require a specific data-generating process and assume that the epidemic unfolds according to the SEIR model up to the point of intervention. It also assumes that overdispersion in transmission is caused by the contact network structure, which may ignore biological mechanisms of overdispersion.^39^ The simultaneous implementation of other NPIs that affect transmission may affect the validity of this model, and more precise modeling should be used to get better sample size estimates for specific settings. In addition, the possibility of imported infections to the trial communities is ignored here, as well as the effect of the intervention on reducing those.

Because of their different required assumptions and parameters and approximations, both the approximation formulae and the simulations should be considered with a range of plausible estimates for the parameters when designing a trial. We propose that investigators considering a cRCT for an NPI follow the following procedure to estimate the required sample size:

1. Calculate approximations to the number of clusters required per arm using the two approximation formulae presented here for likely parameters in their setting. If these are well beyond the point of feasibility for the study, the desired power may not be achievable. If any parameters can be manipulated (e.g., by only enrolling high-incidence clusters or changing the cluster size for implementation), consider other combinations that may reduce the required sample size.
2. Consult the simulation results in Tables S1–S17 to find the parameter combination most similar or combinations which bound the likely parameters for the setting of interest. Extrapolate an estimated sample size from these results and again evaluate the feasibility of this sample size.

3.Conduct a simulation study using the best estimates for the transmission dynamics of the setting of interest using the sample size estimated in steps 1–2 and the planned analysis method. Determine if the empirical power from this simulation study approximately matches the desired power.

In addition to changing test statistics, other methods may be used to reduce the sample size required to achieve the desired power. Increasing the time between intervention and evaluation can increase the power to some extent, although this may make the trial more logistically challenging and make interpretation of effect estimates more challenging. Matching and stratification on cluster-level variables may reduce the variability of results and improve power, again changing the interpretation of estimates.^20, 38, 44^ If the number of clusters are limited but a large number of tests are available, repeated cross-sectional testing may also improve power; this design also allows investigation of time-varying effects.^20^

Further work is required to improve the sizing of large-scale cRCTs in outbreak settings. In particular, analysis of data on the variability of infections at different time points during outbreaks among relevant clusters would enable validation of the assumptions made in these approaches. This data validation would improve both the closed-form approximations used here and the validity of simulations conducted to assess power and sample size. In addition, understanding the variability in *R*_*t*_ across clusters, and covariates or data that can be used to predict *R*_*t*_ in a given cluster, will enable a better understanding of the mechanism of effects of NPIs on transmission. This improved understanding of the estimand will improve sample size and power calculations and potentially point to more efficient estimators.

Randomized trials are key to achieving valid hypothesis tests of the effect of interventions in infectious disease outbreaks. Cluster randomized trials can be used to test the total effect of non-pharmaceutical interventions by comparing the infection trajectory in intervention communities to that in control communities. This analysis demonstrates that in some cases, reasonable power to detect meaningful effect sizes can be achieved for such trials, and it provides investigators with tools to estimate the sample size required.

## Data Availability

The simulated data, code, and results that support the findings of this study are openly available in GitHub at http://www.github.com/jsheen/NPI

## 4.1 Acknowledgements

CJEM and JAH were supported by the Princeton Catalysis Initiative.

## Notes

### Competing Interest Statement

The authors have declared no competing interest.

