## Supporting Information for "The required size of cluster randomized trials of non-pharmaceutical interventions in epidemic settings"

July 2021

### Appendix 1. Approximate Variance Derivations

#### A. Approximation Under Full Testing

Suppose there are $N_1$ intervention and $N_0$ control clusters. For simplicity, we assume that each cluster has $n$ individuals. At time $t$ (immediately before the intervention occurs) and $t+1$ (one generation interval after beginning the intervention), we sample $m_1$ individuals from each cluster and record whether they test positive. Denote the number of individuals who test positive in cluster $j$ at times $t$ and $t+1$ by $Y_{j,t}$ and $Y_{j,t+1}$, respectively. Let $I_t$ and $I_{t+1}$ be random variables representing the proportion of individuals infected at times $t$ and $t+1$, respectively, in a randomly-chosen cluster.

Assume that each cluster has time-varying effective reproductive number $R_t^i$ at time $t$ (looking forward) and overdispersion parameter $k^i$, where $i = 0$ for clusters in the control arm and $i = 1$ for clusters in the intervention arm. Then $I_{t+1}$ and $I_t$ are related by:

$$
\begin{aligned}
I_{j,t+1}|I_{j,t} &= \frac{1}{n}\sum_{\ell=1}^{nI_{j,t}} Q_{j,\ell}, \text{ where} \\
Q_{j,\ell} &\sim NegBin(R_t^i, k^i) \ \forall \ \ell.
\end{aligned}
$$

First, assume that we test the full population within each cluster at both time $t$ and $t+1$, so that $m_1 = n$. Then $Y_{j,t+1} = nI_{j,t+1}$ and $Y_{j,t} = nI_{j,t}$. For each cluster $j$ we can then calculate the expectation and approximate the variance of the statistic $\frac{Y_{j,t+1}}{Y_{j,t}}$. For any cluster $j$ in arm $i$:

$$
\begin{aligned}
E[I_{j,t+1}|I_{j,t}] &= I_t R_t^i. \\
Var(I_{j,t+1}|I_{j,t}) &= \frac{I_t}{n} R_t^i \left(1 + \frac{R_t^i}{k^i}\right). \\
E[Y_{j,t+1}|Y_{j,t}] &= nE\left[I_{j,t+1}|I_{j,t} = \frac{Y_{j,t}}{n}\right] \\
&= n\frac{Y_{j,t}}{n} R_t^i = Y_{j,t} R_t^i. \\
Var(Y_{j,t+1}|Y_{j,t}) &= n^2 Var\left(I_{j,t+1}|I_{j,t} = \frac{Y_{j,t}}{n}\right) \\
&= n^2 \frac{Y_{j,t}}{n^2} R_t^i \left(1 + \frac{R_t^i}{k^i}\right) \\
&= Y_{j,t} R_t^i \left(1 + \frac{R_t^i}{k^i}\right).
\end{aligned}
$$

Now, turning to the test statistic:

$$
\begin{aligned}
E\left[\frac{Y_{j,t+1}}{Y_{j,t}}\right] &= E\left[E\left[\frac{Y_{j,t+1}}{Y_{j,t}}|Y_{j,t}\right]\right] \\
&= E\left[\frac{Y_{j,t}R_t^i}{Y_{j,t}}\right] = E[R_t^i]. \\
Var\left(\frac{Y_{j,t+1}}{Y_{j,t}}\right) &= E\left[Var\left(\frac{Y_{j,t+1}}{Y_{j,t}}|Y_{j,t}\right)\right] + Var\left(E\left[\frac{Y_{j,t+1}}{Y_{j,t}}|Y_{j,t}\right]\right) \\
&= E\left[\frac{R_t^i(1+R_t^i/k^i)}{Y_{j,t}}\right] + Var(R_t^i) \\
&= R_t^i\left(1+\frac{R_t^i}{k^i}\right)E\left[\frac{1}{Y_{j,t}}\right],
\end{aligned}
$$

under the assumption that $R_t^i$ and $k^i$ are equal across clusters within arm $i$ (but may vary across arms). Now, using a Taylor series for $\frac{1}{Y_{j,t}}$ around $\frac{1}{E[Y_{j,t}]}$, we can approximate $E\left[\frac{1}{Y_{j,t}}\right]$ by:

$$
\begin{aligned}
E\left[\frac{1}{Y_{j,t}}\right] &= E\left[\frac{1}{E[Y_{j,t}]} - \frac{Y_{j,t}-E[Y_{j,t}]}{E[Y_{j,t}]^2} + \frac{(Y_{j,t}-E[Y_{j,t}])^2}{E[Y_{j,t}]^3} - \cdots\right] \\
&= \frac{1}{E[Y_{j,t}]} - 0 + \frac{Var(Y_{j,t})}{E[Y_{j,t}]^3} - \cdots
\end{aligned}
$$

And thus we can bound the expectation $E\left[\frac{1}{Y_{j,t}}\right]$ by:

$$
\frac{1}{E[Y_{j,t}]} \le E\left[\frac{1}{Y_{j,t}}\right] \le \frac{1}{E[Y_{j,t}]} + \frac{Var(Y_{j,t})}{E[Y_{j,t}]^3}.
$$

So we can bound the variance in each cluster $j$ in arm $i$ by:

$$
\begin{aligned}
R_t^i\left(1+\frac{R_t^i}{k^i}\right)\frac{1}{E[Y_{j,t}]} &\le Var\left(\frac{Y_{j,t+1}}{Y_{j,t}}\right) \\
&\le R_t^i\left(1+\frac{R_t^i}{k^i}\right)\left(\frac{1}{E[Y_{j,t}]} + \frac{Var(Y_{j,t})}{E[Y_{j,t}]^3}\right). \qquad (1)
\end{aligned}
$$

Equivalently, using $I_{j,t}$ and $I_{j,t+1}$:

$$
\begin{aligned}
R_t^i\left(1+\frac{R_t^i}{k^i}\right)\frac{1}{nE[I_{j,t}]} &\le Var\left(\frac{Y_{j,t+1}}{Y_{j,t}}\right) \\
&\le R_t^i\left(1+\frac{R_t^i}{k^i}\right)\left(\frac{1}{nE[I_{j,t}]} + \frac{Var(I_{j,t})}{nE[I_{j,t}]^3}\right). \qquad (2)
\end{aligned}
$$

#### B. Approximate Variance Accounting for Sampling

Here we use the same notation and epidemic spread assumptions as the previous section, but we now take $m_1$ to be small relative to $n$ (i.e., we have random sampling and ignore any finite population correction).

For any cluster $j$ in intervention arm $i$, we can find the mean and variance of $Y_{j,t+1}$:

$$\begin{aligned}
Y_{j,t+1}|I_{j,t+1} &\sim Bin(m, I_{j,t+1}). \\
E[Y_{j,t+1}|I_{j,t}] &= E\left[E[Y_{j,t+1}|I_{j,t}, I_{j,t+1}]|I_{j,t}\right] \\
&= E[m_1 I_{j,t+1}|I_{j,t}] = m_1 E\left[\frac{1}{n}\sum_{\ell=1}^{nI_{j,t}} Q_{j,\ell}|I_{j,t}\right] \\
&= m_1 \frac{1}{n}(nI_{j,t})R_t^i = m_1 I_{j,t} R_t^i.
\end{aligned}$$

$$\begin{aligned}
Var(Y_{j,t+1}|I_{j,t}) &= E\left[Var(Y_{j,t+1}|I_{j,t+1})|I_{j,t}\right] + Var\left(E[Y_{j,t+1}|I_{j,t+1}]|I_{j,t}\right) \\
&= E\left[m_1 I_{j,t+1}(1 - I_{j,t+1})|I_{j,t}\right] + Var\left(m_1 I_{j,t+1}|I_{j,t}\right) \\
&= m_1\left(E[I_{j,t+1}|I_{j,t}] - E[I_{j,t+1}^2|I_{j,t}]\right) + m_1^2 Var(I_{j,t+1}|I_{j,t}) \\
&= m_1\left(E[I_{j,t+1}|I_{j,t}] - Var(I_{j,t+1}|I_{j,t}) - E[I_{j,t+1}|I_{j,t}]^2\right) + m_1^2 Var(I_{j,t+1}|I_{j,t}) \\
&= m_1 E[I_{j,t+1}|I_{j,t}](1 - E[I_{j,t+1}|I_{j,t}]) + m_1(m_1 - 1)Var(I_{j,t+1}|I_{j,t}) \\
&= m_1 I_{j,t} R_t^i (1 - I_{j,t} R_t^i) + m_1(m_1 - 1)\frac{I_{j,t}}{n} R_t^i (1 + R_t^i/k^i).
\end{aligned}$$

$$\begin{aligned}
E[Y_{j,t+1}] &= m_1 R_t^i E[I_{j,t}]. \\
Var(Y_{j,t+1}) &= Var\left(E[Y_{j,t+1}|I_{j,t}]\right) + E\left[Var(Y_{j,t+1}|I_{j,t})\right] \\
&= m_1^2 (R_t^i)^2 Var(I_{j,t}) + m_1 R_t^i E[I_{j,t}(1 - I_{j,t}R_t^i)] + \frac{m_1(m_1-1)R_t^i(1 + R_t^i/k^i)}{n} E[I_{j,t}] \\
&= m_1 R_t^i \left[(m_1 - 1)R_t^i Var(I_{j,t}) - R_t^i E[I_{j,t}]^2 + E[I_{j,t}]\left(1 + \frac{m_1 - 1}{n}\left[1 + \frac{R_t^i}{k^i}\right]\right)\right].
\end{aligned}$$

Suppose that we sampled everyone in the cluster at time $t$. Then we could use the statistic $\frac{Y_{j,t+1}/m_1}{I_{j,t}}$ to estimate $R_t^i$ in cluster $j$ in arm $i$.

$$\begin{aligned}
E\left[\frac{Y_{j,t+1}}{m_1 I_{j,t}}\right] &= E\left[\frac{E[Y_{j,t+1}|I_{j,t}]}{m_1 I_{j,t}}\right] = E\left[R_t^i\right] = R_t^i. \\
Var\left(\frac{Y_{j,t+1}}{m_1 I_{j,t}}\right) &= E\left[\frac{Var(Y_{j,t+1}|I_{j,t})}{m_1^2 I_{j,t}^2}\right] + Var\left(\frac{E[Y_{j,t+1}|I_{j,t}]}{m_1 I_{j,t}}\right) \\
&= E\left[\frac{R_t^i(1 - I_{j,t}R_t^i)}{m_1 I_{j,t}} + \frac{m_1 - 1}{m_1}\frac{1}{nI_{j,t}} R_t^i(1 + R_t^i/k^i)\right] + Var(R_t^i) \\
&= \frac{R_t^i}{m_1} E\left[\frac{1}{I_{j,t}}\right] - \frac{(R_t^i)^2}{m_1} + \frac{m_1 - 1}{m_1}\frac{R_t^i(1 + R_t^i/k)}{n} E\left[\frac{1}{I_{j,t}}\right].
\end{aligned}$$

Similar to the previous section, by a Taylor series, we can bound $E\left[\frac{1}{I_{j,t}}\right]$:

$$\frac{1}{E[I_{j,t}]} \leq E\left[\frac{1}{I_{j,t}}\right] \leq \frac{1}{E[I_{j,t}]} + \frac{Var(I_{j,t})}{E[I_{j,t}]^3}.$$

So we can bound the variance for any cluster $j$ in intervention arm $i$ by:

$$\begin{aligned}
&\frac{R_t^i}{m_1}\left\{\left[1+\frac{m_1-1}{n}\left(1+\frac{R_t^i}{k^i}\right)\right]\left[\frac{1}{E[I_{j,t}]}\right]-R_t^i\right\} \\
&\quad\leq Var\left(\frac{Y_{j,t+1}}{m_1 I_{j,t}}\right) \\
&\qquad\leq \frac{R_t^i}{m_1}\left\{\left[1+\frac{m_1-1}{n}\left(1+\frac{R_t^i}{k^i}\right)\right]\left[\frac{1}{E[I_{j,t}]}+\frac{Var(I_{j,t})}{E[I_{j,t}]^3}\right]-R_t^i\right\}. \qquad (3)
\end{aligned}$$

In fact, we will sample $m_0$ people in each cluster at time $t$ and can denote the number of individuals who test positive at time $t$ in cluster $j$ by $Y_{j,t}$. Again assuming $m_0$ is small relative to $n$, $Y_{j,t} \sim Bin(m_0, I_{j,t})$ and so $Y_{j,t}/m_0$ estimates $I_{j,t}$.

Conditional on $I_t$, $Y_{j,t}$ has expectation $m_1 I_{j,t}$ and variance $m_1 I_{j,t}(1-I_{j,t})$. So the estimator $\frac{Y_{j,t+1}/m_1}{Y_{j,t}/m_0}$ approximately has expectation $R_t^i$ and has variance greater than that given above.

### Appendix 2. Simulation Algorithms

**Algorithm 1:** Create Simulations For a Combination of Parameters

**Result:** List of simulations with and without intervention, $L_1, L_0$
Input $n, E[I_t], R_0, \Delta, k, I_0$
Initialize $L_\beta, L_d, L_0, L_1, P \leftarrow list()$
**Solve for transmission rate,** $\beta$:
**for** 1 to 500 **do**
    Create graph, $G$, overdispersed by $k$, size $n$
    Initialize transmission rate, $b \leftarrow 0.04$
    **while** $R_0$ of $G$ with $b \geq R_0$ **do**
        $b \leftarrow b - 0.0001$
    Add $b$ to $L_\beta$
$\beta \leftarrow$ median($L_\beta$)
**Solve for day of intervention,** $t$:
**for** 1 to 1000 **do**
    Create graph, $G$, overdispersed by $k$, size $n$
    Run simulation, $s$, for 200 days, transmission $\beta$, initial prevalence $I_0$
    $I(d) \leftarrow$ prevalence of infectious persons, $I$, for each day, $d$, of $s$
    Add $I(d)$ to $L_d$
$t \leftarrow$ min $d|\langle I(d)$ in $L_d | I(d) \neq 0\rangle \geq E[I_t]$ and $|I(d)$ in $L_d| \geq 100$
**Create 3000 simulations**:
**for** 1 to 3000 **do**
    Create graph, $G$ overdispersed by $k$, size $n$
    Run simulation until $t$, transmission $\beta$, initial prevalence $I_0$
    Run simulation for one generation with transmission rate, $\beta$, and with transmission rate, (1-$\Delta$) $\times$ $\beta$
    Add simulation with transmission rate, $\beta$, and with transmission rate,(1-$\Delta$) $\times$ $\beta$ to $L_0, L_1$, respectively
**return** $L_0$, $L_1$

**Algorithm 2:** Find Sufficient Number of Clusters Per Arm, $C_S$, for Two-Sample Welch's t-test, One Generation after Intervention

**Result:** $C_S$ when $\alpha = 5\%$, power $\approx 80\%$
Input $L_0, L_1, m$
Initialize $P \leftarrow list()$
Initialize $C_{min} \leftarrow 1, C_{max} \leftarrow 1000$
**Binary Search**($L_1$, $L_0$,$C_{min}$,$C_{max}$,$m$):
$C \leftarrow$ ceiling($\frac{C_{min}+C_{max}}{2}$)
**for** 1 to 10000 **do**
    Randomly choose $C$ clusters from $L_0$ and $C$ clusters from $L_1$
    Sample $m$ from each cluster at time of intervention, $t$
    $Y_t \leftarrow$ number of sampled infectious individuals at $t$
    Sample $m$ from each cluster one generation after intervention
    $Y_{t+1} \leftarrow$ number of sampled infectious individuals at $t+1$
    Add p-value of two-sample Welch's t-test of $log((Y_{t+1}+1)/(Y_t+1))$ to $P$
Calculate power of $C$, $C_p \leftarrow$ proportion of $P$ with p-value $\leq 0.05$
**if** $C_{max} - C_{min} = 1$ or $C_p$within 0.5% of 80% **then**
    **return** $C$
**else**
    **if** $C_p < 80\%$ **then**
        $C_{max} \leftarrow C$
    **else**
        $C_{min} \leftarrow C$
    **Binary Search**($L_1$, $L_0$,$C_{min}$,$C_{max}$,$m$)

Note: The binary search of Algorithm 2 is terminated if the power at first iteration (which tests 501 clusters per arm) is less than 40% or if the power at second iteration (which tests 751 clusters per arm) is less than 60%. We assume that if either of these conditions are true, Algorithm 2 will not be able to solve for a sufficient number of clusters, $C_S$, $\leq 1000$.

#### Stability of Simulation Algorithm

Solving for the required sample size first requires that we create a stable set of 3000 simulations for each parameter combination, which is done with Algorithm 1. Algorithm 1 is made up of three parts: (1) solve for the transmission rate, (2) solve for the day of intervention, and (3) create 3000 simulations using the solved for transmission rate and day of intervention. To check the stability of (1) we plot the distribution of absolute differences in $\beta$ across two reruns in Figure S1A and find that the average absolute difference across all parameter combinations is $9.7 \times 10^{-5}$ (less than one step of the algorithm, $1 \times 10^{-4}$). To check the stability of (2) we plot the distribution of absolute differences in $t$ across two reruns in Figure S1B and find that the average absolute difference across all parameter combinations is 1.41 days. We have assumed that the 3000 simulations of (3) capture the majority of the possibilities that a single recruited cluster could experience. More specifically, these 3000 simulations are assumed to account for variance in creation of the cluster's contact network, as well as variance in the transmission of the epidemic through a given cluster's contact network.

Algorithm 2, will return the first number of clusters it encounters within 0.5% of 80% (i.e. a window of 0.5%), or will return the result when the difference between the minimum and maximum clusters bounds of the recursive call is 1. For a given tested sample size, the power is assumed to have very small variance. This may lead to a suboptimal sample size if (1) the window of 0.5% is too wide, leading the algorithm to return the first cluster size it encounters within this window when there is a smaller cluster size that also satisfies this window, or (2) if the window of 0.5% is too narrow compared to the variance in the power for a given sample size (i.e. the variance is not small), allowing for the possibility for Algorithm 2 to miss the optimal sample size stochastically. (2) will potentially lead to a solved for number of clusters that is outside of the 0.5% window. We assume the window of 0.5% is narrow enough to be stable against (1). We plot the distribution of power values for parameter sets (two-sample Welch's t-test, one generation after sampling; one-sample t-test on differences when matching on susceptible individuals; one-sample t-test on differences when matching on non-infectious individuals) with solved number of clusters from Algorithm 2 in Figure S1C and find 106/110 parameter sets are within the 0.5% window of 80% power. Furthermore, for those 4 parameter sets outside the 0.5% window, the maximum difference from 80% is 1.1%. Both points show stability against (2).

### Appendix 3. Supplementary Tables and Figures

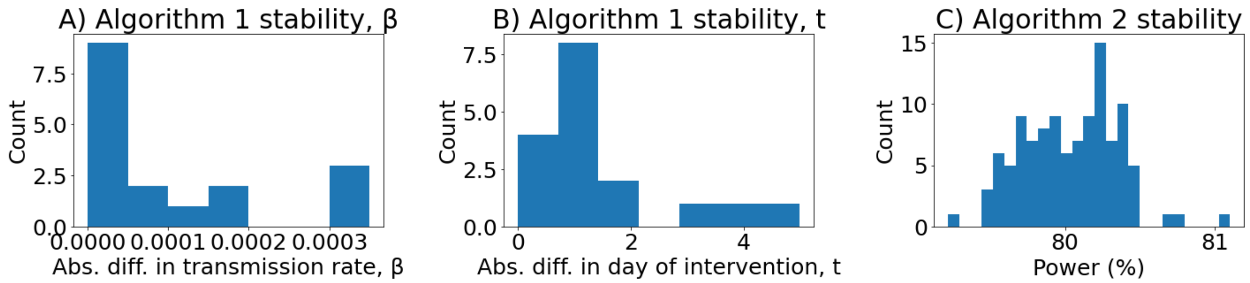


Figure S1: **Distributions from stability analysis of algorithms 1 and 2**. (**A**) Distribution of absolute differences in transmission rates between two runs of Algorithm 1 for all parameter combinations. (**B**) Distribution of absolute differences in days of intervention between two runs of Algorithm 1 for all parameter combinations. (**C**) Distribution of powers for parameter sets with solved number of clusters from Algorithm 2.

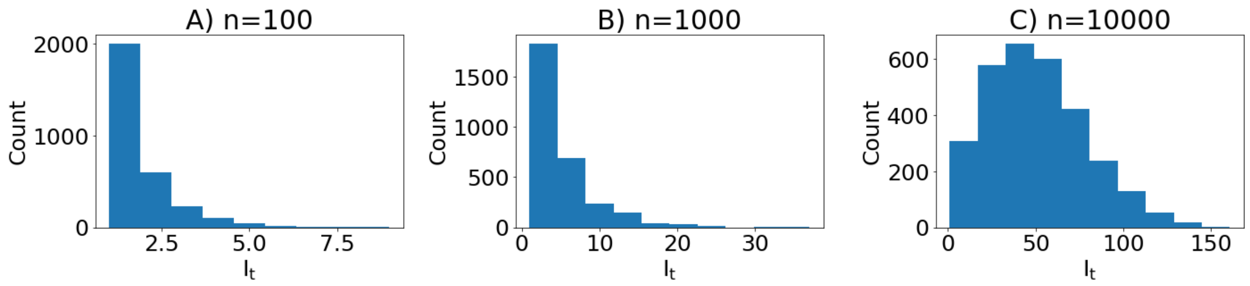


Figure S2: **The distribution of the number of infectious individuals per cluster at time of intervention has large variance**. (**A**) $n = 100$. (**B**) $n = 1000$. (**C**) $n = 10000$. 3000 simulations. $R_0 = 1.5$, $k = 0.7$, targeted $E[I_t] = 2\%$ for $n = 100$ and $E[I_t] = 0.5\%$ for $n = 1000, 10000$. This variance is accounted for in the simulations but not in the approximations.

| $R_0$ | $k$ | $\Delta$ | $t$ | $E[R_t^0]$ | $Var[R_t^0]$ | $E[R_t^1]$ | $Var[R_t^1]$ | $E[I_t]$ | $Var[I_t]$ | $E[I_{t+1}^0]$ | $Var[I_{t+1}^0]$ | $E[I_{t+1}^1]$ | $Var[I_{t+1}^1]$ | $m$ | $C_S$ |
|---|---|---|---|---|---|---|---|---|---|---|---|---|---|---|---|
| 1.5 | 0.7 | 0.2 | 11 | 1.13 | 1.87 | 0.91 | 1.32 | 0.018 | 1.51E-04 | 0.02 | 4.98E-04 | 0.016 | 3.80E-04 | 10 | $\geq 1000$ |
| | | | | | | | | | | | | | | 50 | 845 |
| | | | | | | | | | | | | | | 100 | 408 |
| | | 0.4 | 9 | 1.06 | 1.74 | 0.72 | 1.04 | 0.016 | 9.58E-05 | 0.017 | 4.08E-04 | 0.011 | 2.47E-04 | 10 | $\geq 1000$ |
| | | | | | | | | | | | | | | 50 | 299 |
| | | | | | | | | | | | | | | 100 | 131 |
| | 0.4 | 0.2 | 9 | 0.94 | 1.67 | 0.78 | 1.27 | 0.015 | 9.28E-05 | 0.015 | 3.93E-04 | 0.012 | 3.06E-04 | 10 | $\geq 1000$ |
| | | | | | | | | | | | | | | 50 | $\geq 1000$ |
| | | | | | | | | | | | | | | 100 | 658 |
| | | 0.4 | 10 | 0.95 | 1.64 | 0.65 | 0.81 | 0.016 | 1.20E-04 | 0.016 | 4.45E-04 | 0.011 | 2.20E-04 | 10 | $\geq 1000$ |
| | | | | | | | | | | | | | | 50 | 361 |
| | | | | | | | | | | | | | | 100 | 166 |
| | 0.1 | 0.2 | 8 | 0.53 | 1.11 | 0.45 | 0.79 | 0.013 | 5.04E-05 | 0.008 | 2.37E-04 | 0.007 | 1.59E-04 | 10 | $\geq 1000$ |
| | | | | | | | | | | | | | | 50 | $\geq 1000$ |
| | | | | | | | | | | | | | | 100 | $\geq 1000$ |
| | | 0.4 | 11 | 0.65 | 1.16 | 0.45 | 0.63 | 0.015 | 1.15E-04 | 0.011 | 2.82E-04 | 0.008 | 1.65E-04 | 10 | $\geq 1000$ |
| | | | | | | | | | | | | | | 50 | 611 |
| | | | | | | | | | | | | | | 100 | 248 |
| 2 | 0.7 | 0.2 | 8 | 1.51 | 2.98 | 1.25 | 2.3 | 0.017 | 1.33E-04 | 0.026 | 7.89E-04 | 0.021 | 6.22E-04 | 10 | $\geq 1000$ |
| | | | | | | | | | | | | | | 50 | 814 |
| | | | | | | | | | | | | | | 100 | 426 |
| | | 0.4 | 8 | 1.53 | 3.02 | 0.95 | 1.42 | 0.017 | 1.24E-04 | 0.026 | 8.16E-04 | 0.016 | 3.92E-04 | 10 | 549 |
| | | | | | | | | | | | | | | 50 | 151 |
| | | | | | | | | | | | | | | 100 | 76 |
| | 0.4 | 0.2 | 7 | 1.21 | 2.53 | 1 | 1.91 | 0.015 | 8.77E-05 | 0.019 | 6.10E-04 | 0.016 | 4.62E-04 | 10 | $\geq 1000$ |
| | | | | | | | | | | | | | | 50 | $\geq 1000$ |
| | | | | | | | | | | | | | | 100 | 564 |
| | | 0.4 | 8 | 1.28 | 2.57 | 0.84 | 1.38 | 0.016 | 1.22E-04 | 0.022 | 6.62E-04 | 0.014 | 3.68E-04 | 10 | 876 |
| | | | | | | | | | | | | | | 50 | 228 |
| | | | | | | | | | | | | | | 100 | 115 |
| | 0.1 | 0.2 | 8 | 0.79 | 1.85 | 0.7 | 1.46 | 0.014 | 8.79E-05 | 0.013 | 4.20E-04 | 0.011 | 3.43E-04 | 10 | $\geq 1000$ |
| | | | | | | | | | | | | | | 50 | $\geq 1000$ |
| | | | | | | | | | | | | | | 100 | $\geq 1000$ |
| | | 0.4 | 9 | 0.86 | 1.81 | 0.58 | 0.94 | 0.015 | 1.24E-04 | 0.014 | 4.38E-04 | 0.01 | 2.68E-04 | 10 | $\geq 1000$ |
| | | | | | | | | | | | | | | 50 | 449 |
| | | | | | | | | | | | | | | 100 | 213 |

Table S1: **Full Simulation Results for $n = 100$, Two-Sample Welch's t-test One Generation After Intervention.** $R_0$ = target $R_0$. $k$ = overdispersion. $\Delta$ = % reduction in transmission. $t$ = day of intervention. $R_t^0 = R_t$ without treatment. $R_t^1 = R_t$ with treatment. $I_t$ = proportion of infectious persons at day of intervention. $I_{t+1}^0$ = proportion of infectious persons at time of sampling without treatment. $I_{t+1}^1$ = proportion of infectious persons at time of sampling with treatment. $m$ = number of persons sampled from each cluster. $C_S$ = number of clusters needed for each arm from simulations.

| $R_0$ | $k$ | $\Delta$ | $t$ | $E[R_t^0]$ | $Var[R_t^0]$ | $E[R_t^1]$ | $Var[R_t^1]$ | $E[I_t]$ | $Var[I_t]$ | $E[I_{t+1}^0]$ | $Var[I_{t+1}^0]$ | $E[I_{t+1}^1]$ | $Var[I_{t+1}^1]$ | $m$ | $C_S$ |
|---|---|---|---|---|---|---|---|---|---|---|---|---|---|---|---|
| 1.5 | 0.7 | 0.2 | 27 | 1.5 | 1.71 | 1.18 | 1.1 | 0.0049 | 1.91E-05 | 0.0067 | 3.89E-05 | 0.0054 | 2.77E-05 | 100 | 969 |
| | | | | | | | | | | | | | | 1000 | 142 |
| | | 0.4 | 26 | 1.47 | 1.61 | 0.96 | 0.84 | 0.0048 | 1.87E-05 | 0.0064 | 3.81E-05 | 0.0042 | 1.83E-05 | 100 | 295 |
| | | | | | | | | | | | | | | 1000 | 43 |
| | 0.4 | 0.2 | 28 | 1.32 | 1.59 | 1.13 | 1.14 | 0.0047 | 1.78E-05 | 0.0057 | 3.02E-05 | 0.0049 | 2.33E-05 | 100 | $\geq$ 1000 |
| | | | | | | | | | | | | | | 1000 | 322 |
| | | 0.4 | 28 | 1.33 | 1.48 | 0.91 | 0.79 | 0.0047 | 1.78E-05 | 0.0057 | 3.04E-05 | 0.004 | 1.54E-05 | 100 | 420 |
| | | | | | | | | | | | | | | 1000 | 63 |
| | 0.1 | 0.2 | 36 | 1.16 | 1.26 | 0.97 | 0.93 | 0.0043 | 1.19E-05 | 0.0043 | 1.46E-05 | 0.0036 | 1.01E-05 | 100 | $\geq$ 1000 |
| | | | | | | | | | | | | | | 1000 | 283 |
| | | 0.4 | 32 | 1.21 | 1.41 | 0.85 | 0.78 | 0.0041 | 1.14E-05 | 0.0044 | 1.51E-05 | 0.0031 | 8.50E-06 | 100 | 611 |
| | | | | | | | | | | | | | | 1000 | 68 |
| 2 | 0.7 | 0.2 | 16 | 2.08 | 3.5 | 1.65 | 2.35 | 0.0046 | 1.73E-05 | 0.009 | 7.55E-05 | 0.0071 | 4.94E-05 | 100 | 626 |
| | | | | | | | | | | | | | | 1000 | 158 |
| | | 0.4 | 18 | 2.1 | 3.21 | 1.29 | 1.32 | 0.0054 | 2.48E-05 | 0.0102 | 8.83E-05 | 0.0065 | 3.96E-05 | 100 | 158 |
| | | | | | | | | | | | | | | 1000 | 36 |
| | 0.4 | 0.2 | 18 | 1.91 | 2.97 | 1.5 | 2.01 | 0.0046 | 1.83E-05 | 0.0082 | 6.44E-05 | 0.0065 | 4.38E-05 | 100 | 658 |
| | | | | | | | | | | | | | | 1000 | 146 |
| | | 0.4 | 19 | 1.92 | 2.92 | 1.2 | 1.15 | 0.0049 | 2.02E-05 | 0.0085 | 6.46E-05 | 0.0055 | 2.98E-05 | 100 | 197 |
| | | | | | | | | | | | | | | 1000 | 42 |
| | 0.1 | 0.2 | 22 | 1.55 | 2.2 | 1.29 | 1.63 | 0.0045 | 1.49E-05 | 0.0065 | 3.27E-05 | 0.0054 | 2.43E-05 | 100 | $\geq$ 1000 |
| | | | | | | | | | | | | | | 1000 | 263 |
| | | 0.4 | 21 | 1.56 | 2.28 | 1.01 | 1.02 | 0.0042 | 1.46E-05 | 0.0061 | 3.09E-05 | 0.0041 | 1.64E-05 | 100 | 330 |
| | | | | | | | | | | | | | | 1000 | 55 |

Table S2: **Full Simulation Results for $n = 1000$, Two-Sample Welch's t-test One Generation After Intervention.** $R_0$ = target $R_0$. $k$ = overdispersion. $\Delta$ = % reduction in transmission. $t$ = day of intervention. $R_t^0 = R_t$ without treatment. $R_t^1 = R_t$ with treatment. $I_t$ = proportion of infectious persons at day of intervention. $I_{t+1}^0$ = proportion of infectious persons at time of sampling without treatment. $I_{t+1}^1$ = proportion of infectious persons at time of sampling with treatment. $m$ = number of persons sampled from each cluster. $C_S$ = number of clusters needed for each arm from simulations.

| $R_0$ | $k$ | $\Delta$ | $t$ | $E[R_t^0]$ | $Var[R_t^0]$ | $E[R_t^1]$ | $Var[R_t^1]$ | $E[I_t]$ | $Var[I_t]$ | $E[I_{t+1}^0]$ | $Var[I_{t+1}^0]$ | $E[I_{t+1}^1]$ | $Var[I_{t+1}^1]$ | $m$ | $C_S$ |
|---|---|---|---|---|---|---|---|---|---|---|---|---|---|---|---|
| 1.5 | 0.7 | 0.2 | 49 | 1.36 | 0.17 | 1.13 | 0.15 | 0.0049 | 7.27E-06 | 0.0063 | 1.03E-05 | 0.0053 | 7.46E-06 | 100 | $\geq$ 1000 |
| | | | | | | | | | | | | | | 1000 | 178 |
| | | 0.4 | 50 | 1.36 | 0.14 | 0.92 | 0.08 | 0.005 | 7.76E-06 | 0.0065 | 1.08E-05 | 0.0044 | 5.44E-06 | 100 | 307 |
| | | | | | | | | | | | | | | 1000 | 41 |
| | 0.4 | 0.2 | 73 | 1.16 | 0.17 | 0.98 | 0.12 | 0.0052 | 7.11E-06 | 0.0056 | 6.16E-06 | 0.0047 | 4.58E-06 | 100 | $\geq$ 1000 |
| | | | | | | | | | | | | | | 1000 | 236 |
| | | 0.4 | 68 | 1.21 | 0.24 | 0.83 | 0.1 | 0.0049 | 7.06E-06 | 0.0054 | 6.71E-06 | 0.0038 | 3.72E-06 | 100 | 470 |
| | | | | | | | | | | | | | | 1000 | 55 |
| 2 | 0.7 | 0.2 | 24 | 2.09 | 0.22 | 1.66 | 0.13 | 0.0051 | 5.62E-06 | 0.0104 | 1.99E-05 | 0.0083 | 1.31E-05 | 100 | 501 |
| | | | | | | | | | | | | | | 1000 | 88 |
| | | 0.4 | 24 | 2.08 | 0.21 | 1.28 | 0.08 | 0.0052 | 6.07E-06 | 0.0104 | 2.03E-05 | 0.0065 | 8.85E-06 | 100 | 126 |
| | | | | | | | | | | | | | | 1000 | 21 |
| | 0.4 | 0.2 | 30 | 1.93 | 0.32 | 1.55 | 0.19 | 0.0051 | 8.26E-06 | 0.0092 | 2.00E-05 | 0.0075 | 1.42E-05 | 100 | 658 |
| | | | | | | | | | | | | | | 1000 | 107 |
| | | 0.4 | 30 | 1.93 | 0.36 | 1.23 | 0.15 | 0.0052 | 8.51E-06 | 0.0094 | 2.05E-05 | 0.0061 | 9.68E-06 | 100 | 162 |
| | | | | | | | | | | | | | | 1000 | 26 |
| | 0.1 | 0.2 | 56 | 1.28 | 0.61 | 1.05 | 0.24 | 0.0044 | 5.28E-06 | 0.0047 | 4.08E-06 | 0.004 | 3.13E-06 | 100 | $\geq$ 1000 |
| | | | | | | | | | | | | | | 1000 | 283 |
| | | 0.4 | 55 | 1.3 | 0.52 | 0.89 | 0.2 | 0.0044 | 5.54E-06 | 0.0048 | 4.38E-06 | 0.0034 | 2.48E-06 | 100 | 568 |
| | | | | | | | | | | | | | | 1000 | 66 |

Table S3: **Full Simulation Results for $n = 10000$, Two-Sample Welch's t-test One Generation After Intervention.** $R_0$ = target $R_0$. $k$ = overdispersion. $\Delta$ = % reduction in transmission. $t$ = day of intervention. $R_t^0 = R_t$ without treatment. $R_t^1 = R_t$ with treatment. $I_t$ = proportion of infectious persons at day of intervention. $I_{t+1}^0$ = proportion of infectious persons at time of sampling without treatment. $I_{t+1}^1$ = proportion of infectious persons at time of sampling with treatment. $m$ = number of persons sampled from each cluster. $C_S$ = number of clusters needed for each arm from simulations.

| $R_0$ | $k$ | $\Delta$ | $E[R_t^0]$ | $Var[R_t^0]$ | $E[R_t^1]$ | $Var[R_t^1]$ | $E[I_t]$ | $Var[I_t]$ | $E[I_{t+1}^0]$ | $Var[I_{t+1}^0]$ | $E[I_{t+1}^1]$ | $Var[I_{t+1}^1]$ | $m$ | $C_S$ |
|---|---|---|---|---|---|---|---|---|---|---|---|---|---|---|
| 1.5 | 0.7 | 0.2 | 1.04 | 1.25 | 0.92 | 1.02 | 0.031 | 5.03E-04 | 0.027 | 5.57E-04 | 0.023 | 4.45E-04 | 10 | $\geq 1000$ |
| | | | | | | | | | | | | | 50 | $\geq 1000$ |
| | | | | | | | | | | | | | 100 | 689 |
| | | 0.4 | 1.06 | 1.24 | 0.77 | 0.75 | 0.031 | 4.84E-04 | 0.027 | 5.26E-04 | 0.02 | 3.75E-04 | 10 | $\geq 1000$ |
| | | | | | | | | | | | | | 50 | 275 |
| | | | | | | | | | | | | | 100 | 123 |
| | 0.4 | 0.2 | 0.98 | 1.05 | 0.86 | 0.88 | 0.03 | 4.33E-04 | 0.024 | 4.43E-04 | 0.021 | 3.76E-04 | 10 | $\geq 1000$ |
| | | | | | | | | | | | | | 50 | $\geq 1000$ |
| | | | | | | | | | | | | | 100 | 592 |
| | | 0.4 | 1.03 | 1.2 | 0.71 | 0.68 | 0.029 | 4.23E-04 | 0.024 | 4.56E-04 | 0.018 | 2.98E-04 | 10 | $\geq 1000$ |
| | | | | | | | | | | | | | 50 | 244 |
| | | | | | | | | | | | | | 100 | 105 |
| | 0.1 | 0.2 | 0.79 | 0.96 | 0.67 | 0.73 | 0.024 | 2.58E-04 | 0.016 | 2.69E-04 | 0.013 | 2.12E-04 | 10 | $\geq 1000$ |
| | | | | | | | | | | | | | 50 | $\geq 1000$ |
| | | | | | | | | | | | | | 100 | 642 |
| | | 0.4 | 0.76 | 0.85 | 0.55 | 0.56 | 0.024 | 2.73E-04 | 0.015 | 2.56E-04 | 0.011 | 1.68E-04 | 10 | $\geq 1000$ |
| | | | | | | | | | | | | | 50 | 494 |
| | | | | | | | | | | | | | 100 | 205 |
| 2 | 0.7 | 0.2 | 1.2 | 1.46 | 1.01 | 0.98 | 0.045 | 9.02E-04 | 0.04 | 8.26E-04 | 0.035 | 6.79E-04 | 10 | $\geq 1000$ |
| | | | | | | | | | | | | | 50 | 751 |
| | | | | | | | | | | | | | 100 | 384 |
| | | 0.4 | 1.23 | 1.38 | 0.88 | 0.76 | 0.044 | 8.53E-04 | 0.041 | 8.06E-04 | 0.03 | 5.36E-04 | 10 | 876 |
| | | | | | | | | | | | | | 50 | 182 |
| | | | | | | | | | | | | | 100 | 92 |
| | 0.4 | 0.2 | 1.18 | 1.51 | 1 | 1.06 | 0.041 | 7.65E-04 | 0.035 | 6.57E-04 | 0.03 | 5.49E-04 | 10 | $\geq 1000$ |
| | | | | | | | | | | | | | 50 | 814 |
| | | | | | | | | | | | | | 100 | 400 |
| | | 0.4 | 1.14 | 1.41 | 0.79 | 0.66 | 0.041 | 7.34E-04 | 0.034 | 6.74E-04 | 0.026 | 4.46E-04 | 10 | $\geq 1000$ |
| | | | | | | | | | | | | | 50 | 220 |
| | | | | | | | | | | | | | 100 | 111 |
| | 0.1 | 0.2 | 0.78 | 0.95 | 0.69 | 0.76 | 0.03 | 3.77E-04 | 0.018 | 3.19E-04 | 0.016 | 2.73E-04 | 10 | $\geq 1000$ |
| | | | | | | | | | | | | | 50 | $\geq 1000$ |
| | | | | | | | | | | | | | 100 | $\geq 1000$ |
| | | 0.4 | 0.78 | 0.96 | 0.61 | 0.63 | 0.03 | 3.83E-04 | 0.018 | 3.08E-04 | 0.014 | 2.33E-04 | 10 | $\geq 1000$ |
| | | | | | | | | | | | | | 50 | 751 |
| | | | | | | | | | | | | | 100 | 330 |

Table S4: **Full Simulation Results for** $n = 100$**, Two-Sample Welch's t-test One Generation After Intervention, Day of Intervention t=28** $R_0$ = target $R_0$. $k$ = overdispersion. $\Delta$ = % reduction in transmission. $R_t^0 = R_t$ without treatment. $R_t^1 = R_t$ with treatment. $I_t$ = number of infectious persons at day of intervention. $I_{t+1}^0$ = number of infectious persons at time of sampling without treatment. $I_{t+1}^1$ = number of infectious persons at time of sampling with treatment. $m$ = number of persons sampled from each cluster. $C_S$ = number of clusters needed for each arm from simulations.

| $R_0$ | $k$ | $\Delta$ | $E[R_t^0]$ | $Var[R_t^0]$ | $E[R_t^1]$ | $Var[R_t^1]$ | $E[I_t]$ | $Var[I_t]$ | $E[I_{t+1}^0]$ | $Var[I_{t+1}^0]$ | $E[I_{t+1}^1]$ | $Var[I_{t+1}^1]$ | $m$ | $C_S$ |
|---|---|---|---|---|---|---|---|---|---|---|---|---|---|---|
| 1.5 | 0.7 | 0.2 | 1.45 | 1.63 | 1.19 | 1.15 | 0.0052 | 2.12E-05 | 0.0069 | 4.17E-05 | 0.0056 | 2.79E-05 | 100 | $\geq$ 1000 |
| | | | | | | | | | | | | | 1000 | 188 |
| | | 0.4 | 1.44 | 1.79 | 0.92 | 0.73 | 0.0051 | 2.13E-05 | 0.0066 | 3.94E-05 | 0.0044 | 1.99E-05 | 100 | 295 |
| | | | | | | | | | | | | | 1000 | 43 |
| | 0.4 | 0.2 | 1.36 | 1.71 | 1.09 | 1.03 | 0.0046 | 1.67E-05 | 0.0057 | 2.96E-05 | 0.0046 | 2.03E-05 | 100 | $\geq$ 1000 |
| | | | | | | | | | | | | | 1000 | 189 |
| | | 0.4 | 1.41 | 1.76 | 0.94 | 0.77 | 0.0046 | 1.64E-05 | 0.0058 | 2.93E-05 | 0.004 | 1.52E-05 | 100 | 384 |
| | | | | | | | | | | | | | 1000 | 54 |
| | 0.1 | 0.2 | 1.22 | 1.51 | 1.01 | 1.13 | 0.0037 | 9.31E-06 | 0.0041 | 1.48E-05 | 0.0034 | 1.16E-05 | 100 | $\geq$ 1000 |
| | | | | | | | | | | | | | 1000 | 259 |
| | | 0.4 | 1.21 | 1.47 | 0.83 | 0.77 | 0.0037 | 9.62E-06 | 0.0041 | 1.54E-05 | 0.0028 | 8.09E-06 | 100 | 568 |
| | | | | | | | | | | | | | 1000 | 72 |
| 2 | 0.7 | 0.2 | 1.98 | 2.11 | 1.6 | 1.52 | 0.0101 | 7.72E-05 | 0.0164 | 1.48E-04 | 0.0135 | 1.07E-04 | 100 | 470 |
| | | | | | | | | | | | | | 1000 | 103 |
| | | 0.4 | 1.93 | 2.2 | 1.23 | 0.87 | 0.0101 | 7.78E-05 | 0.0162 | 1.51E-04 | 0.0108 | 7.75E-05 | 100 | 123 |
| | | | | | | | | | | | | | 1000 | 25 |
| | 0.4 | 0.2 | 1.78 | 2.06 | 1.45 | 1.3 | 0.0085 | 5.54E-05 | 0.0126 | 9.28E-05 | 0.0104 | 6.85E-05 | 100 | 642 |
| | | | | | | | | | | | | | 1000 | 136 |
| | | 0.4 | 1.78 | 1.98 | 1.15 | 0.88 | 0.0083 | 5.36E-05 | 0.0123 | 9.17E-05 | 0.0083 | 4.74E-05 | 100 | 169 |
| | | | | | | | | | | | | | 1000 | 31 |
| | 0.1 | 0.2 | 1.55 | 1.7 | 1.28 | 1.3 | 0.0062 | 2.52E-05 | 0.0079 | 3.48E-05 | 0.0066 | 2.67E-05 | 100 | $\geq$ 1000 |
| | | | | | | | | | | | | | 1000 | 174 |
| | | 0.4 | 1.59 | 2.12 | 1.04 | 1 | 0.0061 | 2.54E-05 | 0.0078 | 3.50E-05 | 0.0053 | 1.94E-05 | 100 | 267 |
| | | | | | | | | | | | | | 1000 | 39 |

Table S5: **Full Simulation Results for** $n = 1000$**, Two-Sample Welch's t-test One Generation After Intervention, Day of Intervention t=28** $R_0$ = target $R_0$. $k$ = overdispersion. $\Delta$ = % reduction in transmission. $R_t^0 = R_t$ without treatment. $R_t^1 = R_t$ with treatment. $I_t$ = number of infectious persons at day of intervention. $I_{t+1}^0$ = number of infectious persons at time of sampling without treatment. $I_{t+1}^1$ = number of infectious persons at time of sampling with treatment. $m$ = number of persons sampled from each cluster. $C_S$ = number of clusters needed for each arm from simulations.

| $R_0$ | $k$ | $\Delta$ | $t$ | $E[R_t^0]$ | $Var[R_t^0]$ | $E[R_t^1]$ | $Var[R_t^1]$ | $E[I_t]$ | $Var[I_t]$ | $E[I_{t+1}^0]$ | $Var[I_{t+1}^0]$ | $E[I_{t+1}^1]$ | $Var[I_{t+1}^1]$ | $m$ | $C_S$ | $C_S^0 - C_S$ | $1 - \frac{C_S}{C_S^0}$, % |
|---|---|---|---|---|---|---|---|---|---|---|---|---|---|---|---|---|---|
| 1.5 | 0.7 | 0.2 | 11 | 1.17 | 2.77 | 0.8 | 1.56 | 0.018 | 1.51E-04 | 0.019 | 5.53E-04 | 0.014 | 3.61E-04 | 10 | $\geq 1000$ | - | - |
| | | | | | | | | | | | | | | 50 | 353 | 492 | 58.22 |
| | | | | | | | | | | | | | | 100 | 182 | 226 | 55.39 |
| | | 0.4 | 9 | 1.15 | 2.83 | 0.52 | 1.03 | 0.016 | 9.58E-05 | 0.017 | 5.21E-04 | 0.008 | 1.88E-04 | 10 | 424 | - | - |
| | | | | | | | | | | | | | | 50 | 99 | 200 | 66.89 |
| | | | | | | | | | | | | | | 100 | 49 | 82 | 62.6 |
| | 0.4 | 0.2 | 9 | 0.93 | 2.45 | 0.69 | 1.51 | 0.015 | 9.28E-05 | 0.014 | 4.37E-04 | 0.011 | 2.83E-04 | 10 | $\geq 1000$ | - | - |
| | | | | | | | | | | | | | | 50 | 736 | - | - |
| | | | | | | | | | | | | | | 100 | 392 | 266 | 40.43 |
| | | 0.4 | 10 | 0.94 | 2.33 | 0.48 | 0.9 | 0.016 | 1.20E-04 | 0.015 | 4.24E-04 | 0.008 | 1.79E-04 | 10 | 720 | - | - |
| | | | | | | | | | | | | | | 50 | 162 | 199 | 55.12 |
| | | | | | | | | | | | | | | 100 | 74 | 92 | 55.42 |
| | 0.1 | 0.2 | 8 | 0.44 | 1.03 | 0.35 | 0.73 | 0.013 | 5.04E-05 | 0.006 | 1.69E-04 | 0.005 | 1.28E-04 | 10 | $\geq 1000$ | - | - |
| | | | | | | | | | | | | | | 50 | $\geq 1000$ | - | - |
| | | | | | | | | | | | | | | 100 | $\geq 1000$ | - | - |
| | | 0.4 | 11 | 0.53 | 1.15 | 0.29 | 0.53 | 0.015 | 1.15E-04 | 0.008 | 1.97E-04 | 0.005 | 1.00E-04 | 10 | $\geq 1000$ | - | - |
| | | | | | | | | | | | | | | 50 | 400 | 211 | 34.53 |
| | | | | | | | | | | | | | | 100 | 138 | 110 | 44.35 |
| 2 | 0.7 | 0.2 | 8 | 1.91 | 5.82 | 1.36 | 3.72 | 0.017 | 1.33E-04 | 0.03 | 1.09E-03 | 0.022 | 7.35E-04 | 10 | 845 | - | - |
| | | | | | | | | | | | | | | 50 | 261 | 553 | 67.94 |
| | | | | | | | | | | | | | | 100 | 166 | 260 | 61.03 |
| | | 0.4 | 8 | 1.92 | 5.9 | 0.9 | 1.9 | 0.017 | 1.24E-04 | 0.03 | 1.03E-03 | 0.015 | 3.93E-04 | 10 | 228 | 321 | 58.47 |
| | | | | | | | | | | | | | | 50 | 68 | 83 | 54.97 |
| | | | | | | | | | | | | | | 100 | 42 | 34 | 44.74 |
| | 0.4 | 0.2 | 7 | 1.46 | 4.64 | 1.06 | 3 | 0.015 | 8.77E-05 | 0.022 | 8.37E-04 | 0.016 | 5.26E-04 | 10 | $\geq 1000$ | - | - |
| | | | | | | | | | | | | | | 50 | 424 | - | - |
| | | | | | | | | | | | | | | 100 | 275 | 289 | 51.24 |
| | | 0.4 | 8 | 1.57 | 4.99 | 0.71 | 1.49 | 0.016 | 1.22E-04 | 0.024 | 8.41E-04 | 0.011 | 2.97E-04 | 10 | 299 | 577 | 65.87 |
| | | | | | | | | | | | | | | 50 | 84 | 144 | 63.16 |
| | | | | | | | | | | | | | | 100 | 49 | 66 | 57.39 |
| | 0.1 | 0.2 | 8 | 0.73 | 1.97 | 0.56 | 1.34 | 0.014 | 8.79E-05 | 0.011 | 3.29E-04 | 0.008 | 2.34E-04 | 10 | $\geq 1000$ | - | - |
| | | | | | | | | | | | | | | 50 | $\geq 1000$ | - | - |
| | | | | | | | | | | | | | | 100 | 580 | - | - |
| | | 0.4 | 9 | 0.76 | 1.96 | 0.41 | 0.79 | 0.015 | 1.24E-04 | 0.012 | 3.27E-04 | 0.007 | 1.55E-04 | 10 | $\geq 1000$ | - | - |
| | | | | | | | | | | | | | | 50 | 281 | 168 | 37.42 |
| | | | | | | | | | | | | | | 100 | 123 | 90 | 42.25 |

Table S6: **Full Simulation Results for $n = 100$, Two-Sample Welch's t-test Two Generations After Intervention.** $R_0$ = target $R_0$. $k$ = overdispersion. $\Delta$ = % reduction in transmission. $t$ = day of intervention. $R_t^0 = R_t$ without treatment. $R_t^1 = R_t$ with treatment. $I_t$ = proportion of infectious persons at day of intervention. $I_{t+1}^0$ = proportion of infectious persons at time of sampling without treatment. $I_{t+1}^1$ = proportion of infectious persons at time of sampling with treatment. $m$ = number of persons sampled from each cluster. $C_S$ = number of clusters needed for each arm from simulations. $C_S^0$ = number of clusters needed from simulations using a two-sample Welch's t-test in Table S1.

| $R_0$ | $k$ | $\Delta$ | $t$ | $E[R_t^0]$ | $Var[R_t^0]$ | $E[R_t^1]$ | $Var[R_t^1]$ | $E[I_t]$ | $Var[I_t]$ | $E[I_{t+1}^0]$ | $Var[I_{t+1}^0]$ | $E[I_{t+1}^1]$ | $Var[I_{t+1}^1]$ | $m$ | $C_S$ | $C_S^0 - C_S$ | $1 - \frac{C_S}{C_S^0}$, % |
|---|---|---|---|---|---|---|---|---|---|---|---|---|---|---|---|---|---|
| 1.5 | 0.7 | 0.2 | 27 | 1.96 | 4.98 | 1.2 | 2.01 | 0.0049 | 1.91E-05 | 0.008 | 5.56E-05 | 0.0053 | 2.81E-05 | 100 | 244 | 725 | 74.82 |
| | | | | | | | | | | | | | | 1000 | 59 | 83 | 58.45 |
| | | 0.4 | 26 | 1.95 | 4.53 | 0.77 | 1.08 | 0.0048 | 1.87E-05 | 0.0078 | 5.41E-05 | 0.0032 | 1.28E-05 | 100 | 72 | 223 | 75.59 |
| | | | | | | | | | | | | | | 1000 | 16 | 27 | 62.79 |
| | 0.4 | 0.2 | 28 | 1.66 | 4.17 | 1.12 | 1.98 | 0.0047 | 1.78E-05 | 0.0064 | 3.70E-05 | 0.0045 | 2.11E-05 | 100 | 392 | - | - |
| | | | | | | | | | | | | | | 1000 | 94 | 228 | 70.81 |
| | | 0.4 | 28 | 1.59 | 3.5 | 0.71 | 1.01 | 0.0047 | 1.78E-05 | 0.0063 | 3.60E-05 | 0.0029 | 1.04E-05 | 100 | 112 | 308 | 73.33 |
| | | | | | | | | | | | | | | 1000 | 23 | 40 | 63.49 |
| | 0.1 | 0.2 | 36 | 1.16 | 2.36 | 0.8 | 1.19 | 0.0043 | 1.19E-05 | 0.0038 | 1.32E-05 | 0.0028 | 7.70E-06 | 100 | 861 | - | - |
| | | | | | | | | | | | | | | 1000 | 126 | 157 | 55.48 |
| | | 0.4 | 32 | 1.27 | 2.72 | 0.58 | 0.83 | 0.0041 | 1.14E-05 | 0.004 | 1.41E-05 | 0.0019 | 5.14E-06 | 100 | 205 | 406 | 66.45 |
| | | | | | | | | | | | | | | 1000 | 28 | 40 | 58.82 |
| 2 | 0.7 | 0.2 | 16 | 3.77 | 15.62 | 2.42 | 7.55 | 0.0046 | 1.73E-05 | 0.0146 | 1.59E-04 | 0.0095 | 7.84E-05 | 100 | 138 | 488 | 77.96 |
| | | | | | | | | | | | | | | 1000 | 64 | 94 | 59.49 |
| | | 0.4 | 18 | 3.77 | 15.89 | 1.37 | 2.23 | 0.0054 | 2.48E-05 | 0.0159 | 1.58E-04 | 0.0064 | 3.72E-05 | 100 | 33 | 125 | 79.11 |
| | | | | | | | | | | | | | | 1000 | 14 | 22 | 61.11 |
| | 0.4 | 0.2 | 18 | 3.2 | 12.7 | 1.99 | 5.41 | 0.0046 | 1.83E-05 | 0.0119 | 1.06E-04 | 0.0078 | 5.67E-05 | 100 | 158 | 500 | 75.99 |
| | | | | | | | | | | | | | | 1000 | 64 | 82 | 56.16 |
| | | 0.4 | 19 | 3.14 | 11.11 | 1.21 | 2.11 | 0.0049 | 2.02E-05 | 0.0122 | 9.99E-05 | 0.0051 | 2.61E-05 | 100 | 45 | 152 | 77.16 |
| | | | | | | | | | | | | | | 1000 | 16 | 26 | 61.9 |
| | 0.1 | 0.2 | 22 | 2.04 | 5.67 | 1.42 | 2.98 | 0.0045 | 1.49E-05 | 0.0073 | 3.70E-05 | 0.0052 | 2.11E-05 | 100 | 376 | - | - |
| | | | | | | | | | | | | | | 1000 | 103 | 160 | 60.84 |
| | | 0.4 | 21 | 2.16 | 6.85 | 0.9 | 1.53 | 0.0042 | 1.46E-05 | 0.0072 | 3.74E-05 | 0.0032 | 1.19E-05 | 100 | 88 | 242 | 73.33 |
| | | | | | | | | | | | | | | 1000 | 21 | 34 | 61.82 |

Table S7: **Full Simulation Results for $n = 1000$, Two-Sample Welch's t-test Two Generations After Intervention.** $R_0$ = target $R_0$. $k$ = overdispersion. $\Delta$ = % reduction in transmission. $t$ = day of intervention. $R_t^0 = R_t$ without treatment. $R_t^1 = R_t$ with treatment. $I_t$ = proportion of infectious persons at day of intervention. $I_{t+1}^0$ = proportion of infectious persons at time of sampling without treatment. $I_{t+1}^1$ = proportion of infectious persons at time of sampling with treatment. $m$ = number of persons sampled from each cluster. $C_S$ = number of clusters needed for each arm from simulations. $C_S^0$ = number of clusters needed for each arm from simulations using a two-sample Welch's t-test in Table S2.

| $R_0$ | $k$ | $\Delta$ | $t$ | $E[R_t^0]$ | $Var[R_t^0]$ | $E[R_t^1]$ | $Var[R_t^1]$ | $E[I_t]$ | $Var[I_t]$ | $E[I_{t+1}^0]$ | $Var[I_{t+1}^0]$ | $E[I_{t+1}^1]$ | $Var[I_{t+1}^1]$ | $m$ | $C_S$ | $C_S^0 - C_S$ | $1 - \frac{C_S}{C_S^0}$, % |
|---|---|---|---|---|---|---|---|---|---|---|---|---|---|---|---|---|---|
| 1.5 | 0.7 | 0.2 | 49 | 1.73 | 0.5 | 1.15 | 0.26 | 0.0049 | 7.27E-06 | 0.0076 | 1.13E-05 | 0.0051 | 6.08E-06 | 100 | 251 | - | - |
| | | | | | | | | | | | | | | 1000 | 39 | 139 | 78.09 |
| | | 0.4 | 50 | 1.71 | 0.48 | 0.72 | 0.13 | 0.005 | 7.76E-06 | 0.0077 | 1.21E-05 | 0.0033 | 2.87E-06 | 100 | 70 | 237 | 77.2 |
| | | | | | | | | | | | | | | 1000 | 11 | 30 | 73.17 |
| | 0.4 | 0.2 | 73 | 1.26 | 0.55 | 0.87 | 0.28 | 0.0052 | 7.11E-06 | 0.0055 | 5.14E-06 | 0.0039 | 2.79E-06 | 100 | 470 | - | - |
| | | | | | | | | | | | | | | 1000 | 61 | 175 | 74.15 |
| | | 0.4 | 68 | 1.38 | 0.91 | 0.6 | 0.14 | 0.0049 | 7.06E-06 | 0.0056 | 5.66E-06 | 0.0026 | 1.57E-06 | 100 | 123 | 347 | 73.83 |
| | | | | | | | | | | | | | | 1000 | 15 | 40 | 72.73 |
| 2 | 0.7 | 0.2 | 24 | 3.7 | 1.21 | 2.35 | 0.49 | 0.0051 | 5.62E-06 | 0.0175 | 3.57E-05 | 0.0113 | 1.81E-05 | 100 | 84 | 417 | 83.23 |
| | | | | | | | | | | | | | | 1000 | 20 | 68 | 77.27 |
| | | 0.4 | 24 | 3.67 | 1.16 | 1.36 | 0.17 | 0.0052 | 6.07E-06 | 0.0175 | 3.71E-05 | 0.0067 | 8.19E-06 | 100 | 23 | 103 | 81.75 |
| | | | | | | | | | | | | | | 1000 | 6 | 15 | 71.43 |
| | 0.4 | 0.2 | 30 | 3.11 | 1.91 | 2.02 | 0.72 | 0.0051 | 8.26E-06 | 0.0134 | 2.52E-05 | 0.009 | 1.43E-05 | 100 | 126 | 532 | 80.85 |
| | | | | | | | | | | | | | | 1000 | 28 | 79 | 73.83 |
| | | 0.4 | 30 | 3.13 | 2.25 | 1.22 | 0.27 | 0.0052 | 8.51E-06 | 0.0137 | 2.44E-05 | 0.0057 | 6.52E-06 | 100 | 33 | 129 | 79.63 |
| | | | | | | | | | | | | | | 1000 | 7 | 19 | 73.08 |
| | 0.1 | 0.2 | 56 | 1.5 | 2.56 | 1 | 0.84 | 0.0044 | 5.28E-06 | 0.0044 | 3.12E-06 | 0.0032 | 1.79E-06 | 100 | 689 | - | - |
| | | | | | | | | | | | | | | 1000 | 103 | 180 | 63.6 |
| | | 0.4 | 55 | 1.54 | 2.55 | 0.67 | 0.33 | 0.0044 | 5.54E-06 | 0.0045 | 3.19E-06 | 0.0022 | 1.02E-06 | 100 | 182 | 386 | 67.96 |
| | | | | | | | | | | | | | | 1000 | 24 | 42 | 63.64 |

Table S8: **Full Simulation Results for** $n = 10000$**, Two-Sample Welch's t-test Two Generations After Intervention.** $R_0$ = target $R_0$. $k$ = overdispersion. $\Delta$ = % reduction in transmission. $t$ = day of intervention. $R_t^0 = R_t$ without treatment. $R_t^1 = R_t$ with treatment. $I_t$ = proportion of infectious persons at day of intervention. $I_{t+1}^0$ = proportion of infectious persons at time of sampling without treatment. $I_{t+1}^1$ = proportion of infectious persons at time of sampling with treatment. $m$ = number of persons sampled from each cluster. $C_S$ = number of clusters needed for each arm from simulations. $C_S^0$ = number of clusters needed for each arm from simulations using a two-sample Welch's t-test in Table S3.

| $R_0$ | $k$ | $\Delta$ | $t$ | $E[R_t^0]$ | $Var[R_t^0]$ | $E[R_t^1]$ | $Var[R_t^1]$ | $E[I_t]$ | $Var[I_t]$ | $E[I_{t+1}^0]$ | $Var[I_{t+1}^0]$ | $E[I_{t+1}^1]$ | $Var[I_{t+1}^1]$ | $m$ | $C_S$ | $C_S^0 - C_S$ | $1 - \frac{C_S}{C_S^0}$, % |
|---|---|---|---|---|---|---|---|---|---|---|---|---|---|---|---|---|---|
| 1.5 | 0.7 | 0.2 | 11 | 0.98 | 2.62 | 0.61 | 1.45 | 0.018 | 1.51E-04 | 0.015 | 4.38E-04 | 0.01 | 2.66E-04 | 10 | $\geq$ 1000 | - | - |
| | | | | | | | | | | | | | | 50 | 297 | 548 | 64.85 |
| | | | | | | | | | | | | | | 100 | 146 | 262 | 64.22 |
| | | 0.4 | 9 | 0.99 | 2.97 | 0.35 | 0.69 | 0.016 | 9.58E-05 | 0.014 | 4.38E-04 | 0.005 | 1.15E-04 | 10 | 408 | - | - |
| | | | | | | | | | | | | | | 50 | 97 | 202 | 67.56 |
| | | | | | | | | | | | | | | 100 | 45 | 86 | 65.65 |
| | 0.4 | 0.2 | 9 | 0.75 | 1.99 | 0.5 | 1.13 | 0.015 | 9.28E-05 | 0.011 | 3.21E-04 | 0.007 | 1.98E-04 | 10 | $\geq$ 1000 | - | - |
| | | | | | | | | | | | | | | 50 | 611 | - | - |
| | | | | | | | | | | | | | | 100 | 299 | 359 | 54.56 |
| | | 0.4 | 10 | 0.75 | 1.93 | 0.3 | 0.53 | 0.016 | 1.20E-04 | 0.011 | 3.13E-04 | 0.005 | 1.02E-04 | 10 | 736 | - | - |
| | | | | | | | | | | | | | | 50 | 158 | 203 | 56.23 |
| | | | | | | | | | | | | | | 100 | 68 | 98 | 59.04 |
| | 0.1 | 0.2 | 8 | 0.28 | 0.63 | 0.23 | 0.46 | 0.013 | 5.04E-05 | 0.004 | 9.32E-05 | 0.003 | 6.95E-05 | 10 | $\geq$ 1000 | - | - |
| | | | | | | | | | | | | | | 50 | $\geq$ 1000 | - | - |
| | | | | | | | | | | | | | | 100 | $\geq$ 1000 | - | - |
| | | 0.4 | 11 | 0.33 | 0.72 | 0.16 | 0.28 | 0.015 | 1.15E-04 | 0.005 | 1.11E-04 | 0.002 | 5.04E-05 | 10 | $\geq$ 1000 | - | - |
| | | | | | | | | | | | | | | 50 | 793 | -182 | -29.79 |
| | | | | | | | | | | | | | | 100 | 236 | 12 | 4.84 |
| 2 | 0.7 | 0.2 | 8 | 1.81 | 6.01 | 1.15 | 3.26 | 0.017 | 1.33E-04 | 0.026 | 8.70E-04 | 0.018 | 5.74E-04 | 10 | 697 | - | - |
| | | | | | | | | | | | | | | 50 | 197 | 617 | 75.8 |
| | | | | | | | | | | | | | | 100 | 123 | 303 | 71.13 |
| | | 0.4 | 8 | 1.75 | 5.84 | 0.69 | 1.58 | 0.017 | 1.24E-04 | 0.026 | 8.48E-04 | 0.011 | 2.95E-04 | 10 | 213 | 336 | 61.2 |
| | | | | | | | | | | | | | | 50 | 60 | 91 | 60.26 |
| | | | | | | | | | | | | | | 100 | 35 | 41 | 53.95 |
| | 0.4 | 0.2 | 7 | 1.28 | 4.28 | 0.89 | 2.52 | 0.015 | 8.77E-05 | 0.018 | 6.36E-04 | 0.013 | 3.99E-04 | 10 | $\geq$ 1000 | - | - |
| | | | | | | | | | | | | | | 50 | 470 | - | - |
| | | | | | | | | | | | | | | 100 | 275 | 289 | 51.24 |
| | | 0.4 | 8 | 1.32 | 4 | 0.52 | 1.18 | 0.016 | 1.22E-04 | 0.019 | 5.82E-04 | 0.008 | 2.09E-04 | 10 | 322 | 554 | 63.24 |
| | | | | | | | | | | | | | | 50 | 82 | 146 | 64.04 |
| | | | | | | | | | | | | | | 100 | 43 | 72 | 62.61 |
| | 0.1 | 0.2 | 8 | 0.46 | 1.07 | 0.37 | 0.85 | 0.014 | 8.79E-05 | 0.007 | 1.63E-04 | 0.005 | 1.28E-04 | 10 | $\geq$ 1000 | - | - |
| | | | | | | | | | | | | | | 50 | $\geq$ 1000 | - | - |
| | | | | | | | | | | | | | | 100 | 775 | - | - |
| | | 0.4 | 9 | 0.48 | 1.13 | 0.25 | 0.44 | 0.015 | 1.24E-04 | 0.007 | 1.74E-04 | 0.004 | 8.49E-05 | 10 | $\geq$ 1000 | - | - |
| | | | | | | | | | | | | | | 50 | 501 | -52 | -11.58 |
| | | | | | | | | | | | | | | 100 | 175 | 38 | 17.84 |

Table S9: **Full Simulation Results for $n = 100$, Two-Sample Welch's t-test Three Generations After Intervention.** $R_0$ = target $R_0$. $k$ = overdispersion. $\Delta$ = % reduction in transmission. $t$ = day of intervention. $R_t^0 = R_t$ without treatment. $R_t^1 = R_t$ with treatment. $I_t$ = proportion of infectious persons at day of intervention. $I_{t+1}^0$ = proportion of infectious persons at time of sampling without treatment. $I_{t+1}^1$ = proportion of infectious persons at time of sampling with treatment. $m$ = number of persons sampled from each cluster. $C_S$ = number of clusters needed for each arm from simulations. $C_S^0$ = number of clusters needed for each arm from simulations using a two-sample Welch's t-test in Table S1.

| $R_0$ | $k$ | $\Delta$ | $t$ | $E[R_t^0]$ | $Var[R_t^0]$ | $E[R_t^1]$ | $Var[R_t^1]$ | $E[I_t]$ | $Var[I_t]$ | $E[I_{t+1}^0]$ | $Var[I_{t+1}^0]$ | $E[I_{t+1}^1]$ | $Var[I_{t+1}^1]$ | $m$ | $C_S$ | $C_S^0 - C_S$ | $1-\frac{C_S}{C_S^0}$, % |
|---|---|---|---|---|---|---|---|---|---|---|---|---|---|---|---|---|---|
| 1.5 | 0.7 | 0.2 | 27 | 2.31 | 8.81 | 1.16 | 2.63 | 0.0049 | 1.91E-05 | 0.0086 | 6.09E-05 | 0.0048 | 2.65E-05 | 100 | 126 | 843 | 87 |
| | | | | | | | | | | | | | | 1000 | 37 | 105 | 73.94 |
| | | 0.4 | 26 | 2.33 | 9.03 | 0.6 | 0.94 | 0.0048 | 1.87E-05 | 0.0085 | 6.08E-05 | 0.0024 | 8.97E-06 | 100 | 43 | 252 | 85.42 |
| | | | | | | | | | | | | | | 1000 | 12 | 31 | 72.09 |
| | 0.4 | 0.2 | 28 | 1.89 | 7.01 | 1.02 | 2.32 | 0.0047 | 1.78E-05 | 0.0066 | 3.88E-05 | 0.0038 | 1.69E-05 | 100 | 193 | - | - |
| | | | | | | | | | | | | | | 1000 | 53 | 269 | 83.54 |
| | | 0.4 | 28 | 1.76 | 6.01 | 0.51 | 0.89 | 0.0047 | 1.78E-05 | 0.0063 | 3.70E-05 | 0.002 | 6.76E-06 | 100 | 68 | 352 | 83.81 |
| | | | | | | | | | | | | | | 1000 | 15 | 48 | 76.19 |
| | 0.1 | 0.2 | 36 | 1.06 | 2.73 | 0.63 | 1.13 | 0.0043 | 1.19E-05 | 0.0032 | 1.15E-05 | 0.002 | 5.93E-06 | 100 | 595 | - | - |
| | | | | | | | | | | | | | | 1000 | 92 | 191 | 67.49 |
| | | 0.4 | 32 | 1.17 | 3.29 | 0.37 | 0.53 | 0.0041 | 1.14E-05 | 0.0034 | 1.19E-05 | 0.0012 | 2.83E-06 | 100 | 170 | 441 | 72.18 |
| | | | | | | | | | | | | | | 1000 | 22 | 46 | 67.65 |
| 2 | 0.7 | 0.2 | 16 | 5.6 | 38.41 | 3.11 | 15.48 | 0.0046 | 1.73E-05 | 0.0189 | 1.89E-04 | 0.0111 | 9.27E-05 | 100 | 70 | 556 | 88.82 |
| | | | | | | | | | | | | | | 1000 | 43 | 115 | 72.78 |
| | | 0.4 | 18 | 5.4 | 40.05 | 1.39 | 3.76 | 0.0054 | 2.48E-05 | 0.0195 | 1.75E-04 | 0.0058 | 3.34E-05 | 100 | 19 | 139 | 87.97 |
| | | | | | | | | | | | | | | 1000 | 10 | 26 | 72.22 |
| | 0.4 | 0.2 | 18 | 4.26 | 26.31 | 2.35 | 10.52 | 0.0046 | 1.83E-05 | 0.0138 | 1.14E-04 | 0.0081 | 5.54E-05 | 100 | 92 | 566 | 86.02 |
| | | | | | | | | | | | | | | 1000 | 46 | 100 | 68.49 |
| | | 0.4 | 19 | 4.25 | 24.94 | 1.13 | 2.77 | 0.0049 | 2.02E-05 | 0.0142 | 1.08E-04 | 0.0043 | 2.11E-05 | 100 | 25 | 172 | 87.31 |
| | | | | | | | | | | | | | | 1000 | 11 | 31 | 73.81 |
| | 0.1 | 0.2 | 22 | 2.21 | 9.39 | 1.37 | 4.07 | 0.0045 | 1.49E-05 | 0.0068 | 3.05E-05 | 0.0045 | 1.81E-05 | 100 | 291 | - | - |
| | | | | | | | | | | | | | | 1000 | 78 | 185 | 70.34 |
| | | 0.4 | 21 | 2.4 | 11.21 | 0.71 | 1.42 | 0.0042 | 1.46E-05 | 0.0068 | 3.28E-05 | 0.0024 | 7.91E-06 | 100 | 65 | 265 | 80.3 |
| | | | | | | | | | | | | | | 1000 | 16 | 39 | 70.91 |

Table S10: **Full Simulation Results for $n = 1000$, Two-Sample Welch's t-test Three Generations After Intervention.** $R_0$ = target $R_0$. $k$ = overdispersion. $\Delta$ = % reduction in transmission. $t$ = day of intervention. $R_t^0 = R_t$ without treatment. $R_t^1 = R_t$ with treatment. $I_t$ = proportion of infectious persons at day of intervention. $I_{t+1}^0$ = proportion of infectious persons at time of sampling without treatment. $I_{t+1}^1$ = proportion of infectious persons at time of sampling with treatment. $m$ = number of persons sampled from each cluster. $C_S$ = number of clusters needed for each arm from simulations. $C_S^0$ = number of clusters needed for each arm from simulations using a two-sample Welch's t-test in Table S2.

| $R_0$ | $k$ | $\Delta$ | $t$ | $E[R_t^0]$ | $Var[R_t^0]$ | $E[R_t^1]$ | $Var[R_t^1]$ | $E[I_t]$ | $Var[I_t]$ | $E[I_{t+1}^0]$ | $Var[I_{t+1}^0]$ | $E[I_{t+1}^1]$ | $Var[I_{t+1}^1]$ | $m$ | $C_S$ | $C_S^0 - C_S$ | $1 - \frac{C_S}{C_S^0}$, % |
|---|---|---|---|---|---|---|---|---|---|---|---|---|---|---|---|---|---|
| 1.5 | 0.7 | 0.2 | 49 | 2.06 | 1.26 | 1.11 | 0.36 | 0.0049 | 7.27E-06 | 0.0084 | 1.05E-05 | 0.0047 | 4.37E-06 | 100 | 115 | - | - |
| | | | | | | | | | | | | | | 1000 | 20 | 158 | 88.76 |
| | | 0.4 | 50 | 2.02 | 1.27 | 0.55 | 0.18 | 0.005 | 7.76E-06 | 0.0085 | 1.07E-05 | 0.0024 | 1.50E-06 | 100 | 37 | 270 | 87.95 |
| | | | | | | | | | | | | | | 1000 | 6 | 35 | 85.37 |
| | 0.4 | 0.2 | 73 | 1.33 | 1.54 | 0.75 | 0.42 | 0.0052 | 7.11E-06 | 0.0051 | 4.38E-06 | 0.003 | 1.75E-06 | 100 | 267 | - | - |
| | | | | | | | | | | | | | | 1000 | 37 | 199 | 84.32 |
| | | 0.4 | 68 | 1.49 | 1.9 | 0.43 | 0.16 | 0.0049 | 7.06E-06 | 0.0053 | 4.59E-06 | 0.0017 | 7.46E-07 | 100 | 76 | 394 | 83.83 |
| | | | | | | | | | | | | | | 1000 | 10 | 45 | 81.82 |
| 2 | 0.7 | 0.2 | 24 | 5.31 | 4.94 | 2.95 | 1.23 | 0.0051 | 5.62E-06 | 0.0233 | 2.89E-05 | 0.0135 | 1.65E-05 | 100 | 41 | 460 | 91.82 |
| | | | | | | | | | | | | | | 1000 | 13 | 75 | 85.23 |
| | | 0.4 | 24 | 5.25 | 4.61 | 1.36 | 0.26 | 0.0052 | 6.07E-06 | 0.0232 | 2.99E-05 | 0.0065 | 6.29E-06 | 100 | 12 | 114 | 90.48 |
| | | | | | | | | | | | | | | 1000 | 4 | 17 | 80.95 |
| | 0.4 | 0.2 | 30 | 4.17 | 8.89 | 2.36 | 2 | 0.0051 | 8.26E-06 | 0.0157 | 1.70E-05 | 0.0094 | 9.95E-06 | 100 | 69 | 589 | 89.51 |
| | | | | | | | | | | | | | | 1000 | 20 | 87 | 81.31 |
| | | 0.4 | 30 | 4.14 | 8.63 | 1.13 | 0.48 | 0.0052 | 8.51E-06 | 0.0159 | 1.60E-05 | 0.0049 | 3.91E-06 | 100 | 19 | 143 | 88.27 |
| | | | | | | | | | | | | | | 1000 | 6 | 20 | 76.92 |
| | 0.1 | 0.2 | 56 | 1.64 | 7.41 | 0.93 | 1.83 | 0.0044 | 5.28E-06 | 0.0037 | 2.80E-06 | 0.0024 | 1.33E-06 | 100 | 549 | - | - |
| | | | | | | | | | | | | | | 1000 | 92 | 191 | 67.49 |
| | | 0.4 | 55 | 1.71 | 7.89 | 0.5 | 0.42 | 0.0044 | 5.54E-06 | 0.0038 | 2.86E-06 | 0.0014 | 5.39E-07 | 100 | 150 | 418 | 73.59 |
| | | | | | | | | | | | | | | 1000 | 22 | 44 | 66.67 |

Table S11: **Full Simulation Results for $n = 10000$, Two-Sample Welch's t-test Three Generations After Intervention.** $R_0$ = target $R_0$. $k$ = overdispersion. $\Delta$ = % reduction in transmission. $t$ = day of intervention. $R_t^0 = R_t$ without treatment. $R_t^1 = R_t$ with treatment. $I_t$ = proportion of infectious persons at day of intervention. $I_{t+1}^0$ = proportion of infectious persons at time of sampling without treatment. $I_{t+1}^1$ = proportion of infectious persons at time of sampling with treatment. $m$ = number of persons sampled from each cluster. $C_S$ = number of clusters needed for each arm from simulations. $C_S^0$ = number of clusters needed for each arm from simulations using a two-sample Welch's t-test in Table S3.

| $R_0$ | $k$ | $\Delta$ | $t$ | $E[R_t^0]$ | $Var[R_t^0]$ | $E[R_t^1]$ | $Var[R_t^1]$ | $E[I_t]$ | $Var[I_t]$ | $E[I_{t+1}^0]$ | $Var[I_{t+1}^0]$ | $E[I_{t+1}^1]$ | $Var[I_{t+1}^1]$ | $m$ | $C_S$ | $C_S^0 - C_S$ | $1 - \frac{C_S}{C_S^0}$, % |
|---|---|---|---|---|---|---|---|---|---|---|---|---|---|---|---|---|---|
| 1.5 | 0.7 | 0.2 | 11 | 1.13 | 1.87 | 0.91 | 1.32 | 0.018 | 1.51E-04 | 0.02 | 4.98E-04 | 0.016 | 3.80E-04 | 10 | $\geq 1000$ | - | - |
| | | | | | | | | | | | | | | 50 | 814 | 31 | 3.67 |
| | | | | | | | | | | | | | | 100 | 357 | 51 | 12.5 |
| | | 0.4 | 9 | 1.06 | 1.74 | 0.72 | 1.04 | 0.016 | 9.58E-05 | 0.017 | 4.08E-04 | 0.011 | 2.47E-04 | 10 | $\geq 1000$ | - | - |
| | | | | | | | | | | | | | | 50 | 283 | 16 | 5.35 |
| | | | | | | | | | | | | | | 100 | 111 | 20 | 15.27 |
| | 0.4 | 0.2 | 9 | 0.94 | 1.67 | 0.78 | 1.27 | 0.015 | 9.28E-05 | 0.015 | 3.93E-04 | 0.012 | 3.06E-04 | 10 | $\geq 1000$ | - | - |
| | | | | | | | | | | | | | | 50 | $\geq 1000$ | - | - |
| | | | | | | | | | | | | | | 100 | 533 | 125 | 19 |
| | | 0.4 | 10 | 0.95 | 1.64 | 0.65 | 0.81 | 0.016 | 1.20E-04 | 0.016 | 4.45E-04 | 0.011 | 2.20E-04 | 10 | $\geq 1000$ | - | - |
| | | | | | | | | | | | | | | 50 | 345 | 16 | 4.43 |
| | | | | | | | | | | | | | | 100 | 143 | 23 | 13.86 |
| | 0.1 | 0.2 | 8 | 0.53 | 1.11 | 0.45 | 0.79 | 0.013 | 5.04E-05 | 0.008 | 2.37E-04 | 0.007 | 1.59E-04 | 10 | $\geq 1000$ | - | - |
| | | | | | | | | | | | | | | 50 | $\geq 1000$ | - | - |
| | | | | | | | | | | | | | | 100 | $\geq 1000$ | - | - |
| | | 0.4 | 11 | 0.65 | 1.16 | 0.45 | 0.63 | 0.015 | 1.15E-04 | 0.011 | 2.82E-04 | 0.008 | 1.65E-04 | 10 | $\geq 1000$ | - | - |
| | | | | | | | | | | | | | | 50 | 533 | 78 | 12.77 |
| | | | | | | | | | | | | | | 100 | 199 | 49 | 19.76 |
| 2 | 0.7 | 0.2 | 8 | 1.51 | 2.98 | 1.25 | 2.3 | 0.017 | 1.33E-04 | 0.026 | 7.89E-04 | 0.021 | 6.22E-04 | 10 | $\geq 1000$ | - | - |
| | | | | | | | | | | | | | | 50 | 775 | 39 | 4.79 |
| | | | | | | | | | | | | | | 100 | 322 | 104 | 24.41 |
| | | 0.4 | 8 | 1.53 | 3.02 | 0.95 | 1.42 | 0.017 | 1.24E-04 | 0.026 | 8.16E-04 | 0.016 | 3.92E-04 | 10 | 501 | 48 | 8.74 |
| | | | | | | | | | | | | | | 50 | 142 | 9 | 5.96 |
| | | | | | | | | | | | | | | 100 | 61 | 15 | 19.74 |
| | 0.4 | 0.2 | 7 | 1.21 | 2.53 | 1 | 1.91 | 0.015 | 8.77E-05 | 0.019 | 6.10E-04 | 0.016 | 4.62E-04 | 10 | $\geq 1000$ | - | - |
| | | | | | | | | | | | | | | 50 | 985 | - | - |
| | | | | | | | | | | | | | | 100 | 376 | 188 | 33.33 |
| | | 0.4 | 8 | 1.28 | 2.57 | 0.84 | 1.38 | 0.016 | 1.22E-04 | 0.022 | 6.62E-04 | 0.014 | 3.68E-04 | 10 | 814 | 62 | 7.08 |
| | | | | | | | | | | | | | | 50 | 220 | 8 | 3.51 |
| | | | | | | | | | | | | | | 100 | 88 | 27 | 23.48 |
| | 0.1 | 0.2 | 8 | 0.79 | 1.85 | 0.7 | 1.46 | 0.014 | 8.79E-05 | 0.013 | 4.20E-04 | 0.011 | 3.43E-04 | 10 | $\geq 1000$ | - | - |
| | | | | | | | | | | | | | | 50 | $\geq 1000$ | - | - |
| | | | | | | | | | | | | | | 100 | $\geq 1000$ | - | - |
| | | 0.4 | 9 | 0.86 | 1.81 | 0.58 | 0.94 | 0.015 | 1.24E-04 | 0.014 | 4.38E-04 | 0.01 | 2.68E-04 | 10 | $\geq 1000$ | - | - |
| | | | | | | | | | | | | | | 50 | 384 | 65 | 14.48 |
| | | | | | | | | | | | | | | 100 | 142 | 71 | 33.33 |

Table S12: **Full Simulation Results for $n = 100$, One-Sample t-test With Matching On Sampled Susceptible Individuals.** $R_0$ = target $R_0$. $k$ = overdispersion. $\Delta$ = % reduction in transmission. $t$ = day of intervention. $R_t^0 = R_t$ without treatment. $R_t^1 = R_t$ with treatment. $I_t$ = proportion of infectious persons at day of intervention. $I_{t+1}^0$ = proportion of infectious persons at time of sampling without treatment. $I_{t+1}^1$ = proportion of infectious persons at time of sampling with treatment. $m$ = number of persons sampled from each cluster. $C_S$ = number of clusters needed for each arm from simulations. $C_S^0$ = number of clusters needed for each arm from simulations using a two-sample Welch's t-test in Table S1.

| $R_0$ | $k$ | $\Delta$ | $t$ | $E[R_t^0]$ | $Var[R_t^0]$ | $E[R_t^1]$ | $Var[R_t^1]$ | $E[I_t]$ | $Var[I_t]$ | $E[I_{t+1}^0]$ | $Var[I_{t+1}^0]$ | $E[I_{t+1}^1]$ | $Var[I_{t+1}^1]$ | $m$ | $C_S$ | $C_S^0 - C_S$ | $1 - \frac{C_S}{C_S^0}$, % |
|---|---|---|---|---|---|---|---|---|---|---|---|---|---|---|---|---|---|
| 1.5 | 0.7 | 0.2 | 27 | 1.5 | 1.71 | 1.18 | 1.1 | 0.0049 | 1.91E-05 | 0.0067 | 3.89E-05 | 0.0054 | 2.77E-05 | 100 | 938 | 31 | 3.2 |
| | | | | | | | | | | | | | | 1000 | 138 | 4 | 2.82 |
| | | 0.4 | 26 | 1.47 | 1.61 | 0.96 | 0.84 | 0.0048 | 1.87E-05 | 0.0064 | 3.81E-05 | 0.0042 | 1.83E-05 | 100 | 283 | 12 | 4.07 |
| | | | | | | | | | | | | | | 1000 | 42 | 1 | 2.33 |
| | 0.4 | 0.2 | 28 | 1.32 | 1.59 | 1.13 | 1.14 | 0.0047 | 1.78E-05 | 0.0057 | 3.02E-05 | 0.0049 | 2.33E-05 | 100 | $\geq$ 1000 | - | - |
| | | | | | | | | | | | | | | 1000 | 319 | 3 | 0.93 |
| | | 0.4 | 28 | 1.33 | 1.48 | 0.91 | 0.79 | 0.0047 | 1.78E-05 | 0.0057 | 3.04E-05 | 0.004 | 1.54E-05 | 100 | 408 | 12 | 2.86 |
| | | | | | | | | | | | | | | 1000 | 62 | 1 | 1.59 |
| | 0.1 | 0.2 | 36 | 1.16 | 1.26 | 0.97 | 0.93 | 0.0043 | 1.19E-05 | 0.0043 | 1.46E-05 | 0.0036 | 1.01E-05 | 100 | $\geq$ 1000 | - | - |
| | | | | | | | | | | | | | | 1000 | 283 | 0 | 0 |
| | | 0.4 | 32 | 1.21 | 1.41 | 0.85 | 0.78 | 0.0041 | 1.14E-05 | 0.0044 | 1.51E-05 | 0.0031 | 8.50E-06 | 100 | 595 | 16 | 2.62 |
| | | | | | | | | | | | | | | 1000 | 70 | -2 | -2.94 |
| 2 | 0.7 | 0.2 | 16 | 2.08 | 3.5 | 1.65 | 2.35 | 0.0046 | 1.73E-05 | 0.009 | 7.55E-05 | 0.0071 | 4.94E-05 | 100 | 619 | 7 | 1.12 |
| | | | | | | | | | | | | | | 1000 | 138 | 20 | 12.66 |
| | | 0.4 | 18 | 2.1 | 3.21 | 1.29 | 1.32 | 0.0054 | 2.48E-05 | 0.0102 | 8.83E-05 | 0.0065 | 3.96E-05 | 100 | 158 | 0 | 0 |
| | | | | | | | | | | | | | | 1000 | 33 | 3 | 8.33 |
| | 0.4 | 0.2 | 18 | 1.91 | 2.97 | 1.5 | 2.01 | 0.0046 | 1.83E-05 | 0.0082 | 6.44E-05 | 0.0065 | 4.38E-05 | 100 | 658 | 0 | 0 |
| | | | | | | | | | | | | | | 1000 | 119 | 27 | 18.49 |
| | | 0.4 | 19 | 1.92 | 2.92 | 1.2 | 1.15 | 0.0049 | 2.02E-05 | 0.0085 | 6.46E-05 | 0.0055 | 2.98E-05 | 100 | 197 | 0 | 0 |
| | | | | | | | | | | | | | | 1000 | 39 | 3 | 7.14 |
| | 0.1 | 0.2 | 22 | 1.55 | 2.2 | 1.29 | 1.63 | 0.0045 | 1.49E-05 | 0.0065 | 3.27E-05 | 0.0054 | 2.43E-05 | 100 | $\geq$ 1000 | - | - |
| | | | | | | | | | | | | | | 1000 | 220 | 43 | 16.35 |
| | | 0.4 | 21 | 1.56 | 2.28 | 1.01 | 1.02 | 0.0042 | 1.46E-05 | 0.0061 | 3.09E-05 | 0.0041 | 1.64E-05 | 100 | 322 | 8 | 2.42 |
| | | | | | | | | | | | | | | 1000 | 45 | 10 | 18.18 |

Table S13: **Full Simulation Results for $n = 1000$, One-Sample t-test With Matching On Sampled Susceptible Individuals.** $R_0$ = target $R_0$. $k$ = overdispersion. $\Delta$ = % reduction in transmission. $t$ = day of intervention. $R_t^0 = R_t$ without treatment. $R_t^1 = R_t$ with treatment. $I_t$ = proportion of infectious persons at day of intervention. $I_{t+1}^0$ = proportion of infectious persons at time of sampling without treatment. $I_{t+1}^1$ = proportion of infectious persons at time of sampling with treatment. $m$ = number of persons sampled from each cluster. $C_S$ = number of clusters needed for each arm from simulations. $C_S^0$ = number of clusters needed for each arm from simulations using a two-sample Welch's t-test in Table S2.

| $R_0$ | $k$ | $\Delta$ | $t$ | $E[R_t^0]$ | $Var[R_t^0]$ | $E[R_t^1]$ | $Var[R_t^1]$ | $E[I_t]$ | $Var[I_t]$ | $E[I_{t+1}^0]$ | $Var[I_{t+1}^0]$ | $E[I_{t+1}^1]$ | $Var[I_{t+1}^1]$ | $m$ | $C_S$ | $C_S^0 - C_S$ | $1 - \frac{C_S}{C_S^0}$, % |
|---|---|---|---|---|---|---|---|---|---|---|---|---|---|---|---|---|---|
| 1.5 | 0.7 | 0.2 | 49 | 1.36 | 0.17 | 1.13 | 0.15 | 0.0049 | 7.27E-06 | 0.0063 | 1.03E-05 | 0.0053 | 7.46E-06 | 100 | $\geq$ 1000 | - | - |
| | | | | | | | | | | | | | | 1000 | 174 | 4 | 2.25 |
| | | 0.4 | 50 | 1.36 | 0.14 | 0.92 | 0.08 | 0.005 | 7.76E-06 | 0.0065 | 1.08E-05 | 0.0044 | 5.44E-06 | 100 | 299 | 8 | 2.61 |
| | | | | | | | | | | | | | | 1000 | 41 | 0 | 0 |
| | 0.4 | 0.2 | 73 | 1.16 | 0.17 | 0.98 | 0.12 | 0.0052 | 7.11E-06 | 0.0056 | 6.16E-06 | 0.0047 | 4.58E-06 | 100 | $\geq$ 1000 | - | - |
| | | | | | | | | | | | | | | 1000 | 228 | 8 | 3.39 |
| | | 0.4 | 68 | 1.21 | 0.24 | 0.83 | 0.1 | 0.0049 | 7.06E-06 | 0.0054 | 6.71E-06 | 0.0038 | 3.72E-06 | 100 | 455 | 15 | 3.19 |
| | | | | | | | | | | | | | | 1000 | 55 | 0 | 0 |
| 2 | 0.7 | 0.2 | 24 | 2.09 | 0.22 | 1.66 | 0.13 | 0.0051 | 5.62E-06 | 0.0104 | 1.99E-05 | 0.0083 | 1.31E-05 | 100 | 494 | 7 | 1.4 |
| | | | | | | | | | | | | | | 1000 | 88 | 0 | 0 |
| | | 0.4 | 24 | 2.08 | 0.21 | 1.28 | 0.08 | 0.0052 | 6.07E-06 | 0.0104 | 2.03E-05 | 0.0065 | 8.85E-06 | 100 | 126 | 0 | 0 |
| | | | | | | | | | | | | | | 1000 | 22 | -1 | -4.76 |
| | 0.4 | 0.2 | 30 | 1.93 | 0.32 | 1.55 | 0.19 | 0.0051 | 8.26E-06 | 0.0092 | 2.00E-05 | 0.0075 | 1.42E-05 | 100 | 626 | 32 | 4.86 |
| | | | | | | | | | | | | | | 1000 | 103 | 4 | 3.74 |
| | | 0.4 | 30 | 1.93 | 0.36 | 1.23 | 0.15 | 0.0052 | 8.51E-06 | 0.0094 | 2.05E-05 | 0.0061 | 9.68E-06 | 100 | 159 | 3 | 1.85 |
| | | | | | | | | | | | | | | 1000 | 27 | -1 | -3.85 |
| | 0.1 | 0.2 | 56 | 1.28 | 0.61 | 1.05 | 0.24 | 0.0044 | 5.28E-06 | 0.0047 | 4.08E-06 | 0.004 | 3.13E-06 | 100 | $\geq$ 1000 | - | - |
| | | | | | | | | | | | | | | 1000 | 251 | 32 | 11.31 |
| | | 0.4 | 55 | 1.3 | 0.52 | 0.89 | 0.2 | 0.0044 | 5.54E-06 | 0.0048 | 4.38E-06 | 0.0034 | 2.48E-06 | 100 | 533 | 35 | 6.16 |
| | | | | | | | | | | | | | | 1000 | 61 | 5 | 7.58 |

Table S14: **Full Simulation Results for $n = 10000$, One-Sample t-test With Matching On Sampled Susceptible Individuals.** $R_0$ = target $R_0$. $k$ = overdispersion. $\Delta$ = % reduction in transmission. $t$ = day of intervention. $R_t^0 = R_t$ without treatment. $R_t^1 = R_t$ with treatment. $I_t$ = proportion of infectious persons at day of intervention. $I_{t+1}^0$ = proportion of infectious persons at time of sampling without treatment. $I_{t+1}^1$ = proportion of infectious persons at time of sampling with treatment. $m$ = number of persons sampled from each cluster. $C_S$ = number of clusters needed for each arm from simulations. $C_S^0$ = number of clusters needed for each arm from simulations using a two-sample Welch's t-test in Table S3.

| $R_0$ | $k$ | $\Delta$ | $t$ | $E[R_t^0]$ | $Var[R_t^0]$ | $E[R_t^1]$ | $Var[R_t^1]$ | $E[I_t]$ | $Var[I_t]$ | $E[I_{t+1}^0]$ | $Var[I_{t+1}^0]$ | $E[I_{t+1}^1]$ | $Var[I_{t+1}^1]$ | $m$ | $C_S$ | $C_S^0 - C_S$ | $1 - \frac{C_S}{C_S^0}$, % | 1 - $\frac{C_S}{C_S^M}$, % |
|---|---|---|---|---|---|---|---|---|---|---|---|---|---|---|---|---|---|---|
| 1.5 | 0.7 | 0.2 | 11 | 1.13 | 1.87 | 0.91 | 1.32 | 0.018 | 1.51E-04 | 0.02 | 4.98E-04 | 0.016 | 3.80E-04 | 10 | $\geq$ 1000 | - | - | - |
| | | | | | | | | | | | | | | 50 | 611 | 234 | 27.69 | 24.94 |
| | | | | | | | | | | | | | | 100 | 408 | 0 | 0 | -14.29 |
| | | 0.4 | 9 | 1.06 | 1.74 | 0.72 | 1.04 | 0.016 | 9.58E-05 | 0.017 | 4.08E-04 | 0.011 | 2.47E-04 | 10 | 720 | - | - | - |
| | | | | | | | | | | | | | | 50 | 205 | 94 | 31.44 | 27.56 |
| | | | | | | | | | | | | | | 100 | 132 | -1 | -0.76 | -18.92 |
| | 0.4 | 0.2 | 9 | 0.94 | 1.67 | 0.78 | 1.27 | 0.015 | 9.28E-05 | 0.015 | 3.93E-04 | 0.012 | 3.06E-04 | 10 | $\geq$ 1000 | - | - | - |
| | | | | | | | | | | | | | | 50 | 969 | - | - | - |
| | | | | | | | | | | | | | | 100 | 626 | 32 | 4.86 | -17.45 |
| | | 0.4 | 10 | 0.95 | 1.64 | 0.65 | 0.81 | 0.016 | 1.20E-04 | 0.016 | 4.45E-04 | 0.011 | 2.20E-04 | 10 | 783 | - | - | - |
| | | | | | | | | | | | | | | 50 | 259 | 102 | 28.25 | 24.93 |
| | | | | | | | | | | | | | | 100 | 169 | -3 | -1.81 | -18.18 |
| | 0.1 | 0.2 | 8 | 0.53 | 1.11 | 0.45 | 0.79 | 0.013 | 5.04E-05 | 0.008 | 2.37E-04 | 0.007 | 1.59E-04 | 10 | $\geq$ 1000 | - | - | - |
| | | | | | | | | | | | | | | 50 | $\geq$ 1000 | - | - | - |
| | | | | | | | | | | | | | | 100 | $\geq$ 1000 | - | - | - |
| | | 0.4 | 11 | 0.65 | 1.16 | 0.45 | 0.63 | 0.015 | 1.15E-04 | 0.011 | 2.82E-04 | 0.008 | 1.65E-04 | 10 | $\geq$ 1000 | - | - | - |
| | | | | | | | | | | | | | | 50 | 388 | 223 | 36.5 | 27.2 |
| | | | | | | | | | | | | | | 100 | 244 | 4 | 1.61 | -22.61 |
| 2 | 0.7 | 0.2 | 8 | 1.51 | 2.98 | 1.25 | 2.3 | 0.017 | 1.33E-04 | 0.026 | 7.89E-04 | 0.021 | 6.22E-04 | 10 | $\geq$ 1000 | - | - | - |
| | | | | | | | | | | | | | | 50 | 642 | 172 | 21.13 | 17.16 |
| | | | | | | | | | | | | | | 100 | 424 | 2 | 0.47 | -31.68 |
| | | 0.4 | 8 | 1.53 | 3.02 | 0.95 | 1.42 | 0.017 | 1.24E-04 | 0.026 | 8.16E-04 | 0.016 | 3.92E-04 | 10 | 349 | 200 | 36.43 | 30.34 |
| | | | | | | | | | | | | | | 50 | 115 | 36 | 23.84 | 19.01 |
| | | | | | | | | | | | | | | 100 | 78 | -2 | -2.63 | -27.87 |
| | 0.4 | 0.2 | 7 | 1.21 | 2.53 | 1 | 1.91 | 0.015 | 8.77E-05 | 0.019 | 6.10E-04 | 0.016 | 4.62E-04 | 10 | $\geq$ 1000 | - | - | - |
| | | | | | | | | | | | | | | 50 | 814 | - | - | 17.36 |
| | | | | | | | | | | | | | | 100 | 549 | 15 | 2.66 | -46.01 |
| | | 0.4 | 8 | 1.28 | 2.57 | 0.84 | 1.38 | 0.016 | 1.22E-04 | 0.022 | 6.62E-04 | 0.014 | 3.68E-04 | 10 | 529 | 347 | 39.61 | 35.01 |
| | | | | | | | | | | | | | | 50 | 170 | 58 | 25.44 | 22.73 |
| | | | | | | | | | | | | | | 100 | 115 | 0 | 0 | -30.68 |
| | 0.1 | 0.2 | 8 | 0.79 | 1.85 | 0.7 | 1.46 | 0.014 | 8.79E-05 | 0.013 | 4.20E-04 | 0.011 | 3.43E-04 | 10 | $\geq$ 1000 | - | - | - |
| | | | | | | | | | | | | | | 50 | $\geq$ 1000 | - | - | - |
| | | | | | | | | | | | | | | 100 | $\geq$ 1000 | - | - | - |
| | | 0.4 | 9 | 0.86 | 1.81 | 0.58 | 0.94 | 0.015 | 1.24E-04 | 0.014 | 4.38E-04 | 0.01 | 2.68E-04 | 10 | 985 | - | - | - |
| | | | | | | | | | | | | | | 50 | 314 | 135 | 30.07 | 18.23 |
| | | | | | | | | | | | | | | 100 | 205 | 8 | 3.76 | -44.37 |

Table S15: **Full Simulation Results for $n = 100$, One-Sample t-test With Matching On Sampled Non-Infectious Individuals.** $R_0 =$ target $R_0$. $k =$ overdispersion. $\Delta =$ % reduction in transmission. $t =$ day of intervention. $R_t^0 = R_t$ without treatment. $R_t^1 = R_t$ with treatment. $I_t =$ proportion of infectious persons at day of intervention. $I_{t+1}^0 =$ proportion of infectious persons at time of sampling without treatment. $I_{t+1}^1 =$ proportion of infectious persons at time of sampling with treatment. $m$ = number of persons sampled from each cluster. $C_S =$ number of clusters needed for each arm from simulations.$C_S^0 =$ number of clusters needed for each arm from simulations using a two-sample Welch's t-test in Table S1. $C_S^M =$ number of clusters needed for each arm from simulations using a two-sample Weclh's t-test in Table S12.

| $R_0$ | $k$ | $\Delta$ | $t$ | $E[R_t^0]$ | $Var[R_t^0]$ | $E[R_t^1]$ | $Var[R_t^1]$ | $E[I_t]$ | $Var[I_t]$ | $E[I_{t+1}^0]$ | $Var[I_{t+1}^0]$ | $E[I_{t+1}^1]$ | $Var[I_{t+1}^1]$ | $m$ | $C_S$ | $C_S^0 - C_S$ | $1 - \frac{C_S}{C_S^0}$, % | 1 - $\frac{C_S}{C_S^M}$, % |
|---|---|---|---|---|---|---|---|---|---|---|---|---|---|---|---|---|---|---|
| 1.5 | 0.7 | 0.2 | 27 | 1.5 | 1.71 | 1.18 | 1.1 | 0.0049 | 1.91E-05 | 0.0067 | 3.89E-05 | 0.0054 | 2.77E-05 | 100 | 689 | 280 | 28.9 | 26.55 |
| | | | | | | | | | | | | | | 1000 | 138 | 4 | 2.82 | 0 |
| | | 0.4 | 26 | 1.47 | 1.61 | 0.96 | 0.84 | 0.0048 | 1.87E-05 | 0.0064 | 3.81E-05 | 0.0042 | 1.83E-05 | 100 | 201 | 94 | 31.86 | 28.98 |
| | | | | | | | | | | | | | | 1000 | 43 | 0 | 0 | -2.38 |
| | 0.4 | 0.2 | 28 | 1.32 | 1.59 | 1.13 | 1.14 | 0.0047 | 1.78E-05 | 0.0057 | 3.02E-05 | 0.0049 | 2.33E-05 | 100 | $\geq$ 1000 | - | - | - |
| | | | | | | | | | | | | | | 1000 | 330 | -8 | -2.48 | -3.45 |
| | | 0.4 | 28 | 1.33 | 1.48 | 0.91 | 0.79 | 0.0047 | 1.78E-05 | 0.0057 | 3.04E-05 | 0.004 | 1.54E-05 | 100 | 275 | 145 | 34.52 | 32.6 |
| | | | | | | | | | | | | | | 1000 | 64 | -1 | -1.59 | -3.23 |
| | 0.1 | 0.2 | 36 | 1.16 | 1.26 | 0.97 | 0.93 | 0.0043 | 1.19E-05 | 0.0043 | 1.46E-05 | 0.0036 | 1.01E-05 | 100 | $\geq$ 1000 | - | - | - |
| | | | | | | | | | | | | | | 1000 | 283 | 0 | 0 | 0 |
| | | 0.4 | 32 | 1.21 | 1.41 | 0.85 | 0.78 | 0.0041 | 1.14E-05 | 0.0044 | 1.51E-05 | 0.0031 | 8.50E-06 | 100 | 361 | 250 | 40.92 | 39.33 |
| | | | | | | | | | | | | | | 1000 | 70 | -2 | -2.94 | 0 |
| 2 | 0.7 | 0.2 | 16 | 2.08 | 3.5 | 1.65 | 2.35 | 0.0046 | 1.73E-05 | 0.009 | 7.55E-05 | 0.0071 | 4.94E-05 | 100 | 494 | 132 | 21.09 | 20.19 |
| | | | | | | | | | | | | | | 1000 | 158 | 0 | 0 | -14.49 |
| | | 0.4 | 18 | 2.1 | 3.21 | 1.29 | 1.32 | 0.0054 | 2.48E-05 | 0.0102 | 8.83E-05 | 0.0065 | 3.96E-05 | 100 | 125 | 33 | 20.89 | 20.89 |
| | | | | | | | | | | | | | | 1000 | 36 | 0 | 0 | -9.09 |
| | 0.4 | 0.2 | 18 | 1.91 | 2.97 | 1.5 | 2.01 | 0.0046 | 1.83E-05 | 0.0082 | 6.44E-05 | 0.0065 | 4.38E-05 | 100 | 517 | 141 | 21.43 | 21.43 |
| | | | | | | | | | | | | | | 1000 | 142 | 4 | 2.74 | -19.33 |
| | | 0.4 | 19 | 1.92 | 2.92 | 1.2 | 1.15 | 0.0049 | 2.02E-05 | 0.0085 | 6.46E-05 | 0.0055 | 2.98E-05 | 100 | 150 | 47 | 23.86 | 23.86 |
| | | | | | | | | | | | | | | 1000 | 42 | 0 | 0 | -7.69 |
| | 0.1 | 0.2 | 22 | 1.55 | 2.2 | 1.29 | 1.63 | 0.0045 | 1.49E-05 | 0.0065 | 3.27E-05 | 0.0054 | 2.43E-05 | 100 | 969 | - | - | - |
| | | | | | | | | | | | | | | 1000 | 251 | 12 | 4.56 | -14.09 |
| | | 0.4 | 21 | 1.56 | 2.28 | 1.01 | 1.02 | 0.0042 | 1.46E-05 | 0.0061 | 3.09E-05 | 0.0041 | 1.64E-05 | 100 | 228 | 102 | 30.91 | 29.19 |
| | | | | | | | | | | | | | | 1000 | 53 | 2 | 3.64 | -17.78 |

Table S16: **Full Simulation Results for $n = 1000$, One-Sample t-test With Matching On Sampled Non-Infectious Individuals.** $R_0$ = target $R_0$. $k$ = overdispersion. $\Delta$ = % reduction in transmission. $t$ = day of intervention. $R_t^0 = R_t$ without treatment. $R_t^1 = R_t$ with treatment. $I_t$ = proportion of infectious persons at day of intervention. $I_{t+1}^0$ = proportion of infectious persons at time of sampling without treatment. $I_{t+1}^1$ = proportion of infectious persons at time of sampling with treatment. $m$ = number of persons sampled from each cluster. $C_S$ = number of clusters needed for each arm from simulations.$C_S^0$ = number of clusters needed for each arm from simulations using a two-sample Welch's t-test in Table S2. $C_S^M$ = number of clusters needed for each arm from simulations using a two-sample Weclh's t-test in Table S13.

| $R_0$ | $k$ | $\Delta$ | $t$ | $E[R_t^0]$ | $Var[R_t^0]$ | $E[R_t^1]$ | $Var[R_t^1]$ | $E[I_t]$ | $Var[I_t]$ | $E[I_{t+1}^0]$ | $Var[I_{t+1}^0]$ | $E[I_{t+1}^1]$ | $Var[I_{t+1}^1]$ | $m$ | $C_S$ | $C_S^0 - C_S$ | $1 - \frac{C_S}{C_S^0}$, % | 1 - $\frac{C_S}{C_S^M}$, % |
|---|---|---|---|---|---|---|---|---|---|---|---|---|---|---|---|---|---|---|
| 1.5 | 0.7 | 0.2 | 49 | 1.36 | 0.17 | 1.13 | 0.15 | 0.0049 | 7.27E-06 | 0.0063 | 1.03E-05 | 0.0053 | 7.46E-06 | 100 | 799 | - | - | - |
| | | | | | | | | | | | | | | 1000 | 142 | 32 | 20.22 | 18.39 |
| | | 0.4 | 50 | 1.36 | 0.14 | 0.92 | 0.08 | 0.005 | 7.76E-06 | 0.0065 | 1.08E-05 | 0.0044 | 5.44E-06 | 100 | 189 | 110 | 38.44 | 36.79 |
| | | | | | | | | | | | | | | 1000 | 33 | 8 | 19.51 | 19.51 |
| | 0.4 | 0.2 | 73 | 1.16 | 0.17 | 0.98 | 0.12 | 0.0052 | 7.11E-06 | 0.0056 | 6.16E-06 | 0.0047 | 4.58E-06 | 100 | $\geq$ 1000 | - | - | - |
| | | | | | | | | | | | | | | 1000 | 178 | 50 | 24.58 | 21.93 |
| | | 0.4 | 68 | 1.21 | 0.24 | 0.83 | 0.1 | 0.0049 | 7.06E-06 | 0.0054 | 6.71E-06 | 0.0038 | 3.72E-06 | 100 | 271 | 184 | 42.34 | 40.44 |
| | | | | | | | | | | | | | | 1000 | 44 | 11 | 20 | 20 |
| 2 | 0.7 | 0.2 | 24 | 2.09 | 0.22 | 1.66 | 0.13 | 0.0051 | 5.62E-06 | 0.0104 | 1.99E-05 | 0.0083 | 1.31E-05 | 100 | 345 | 149 | 31.14 | 30.16 |
| | | | | | | | | | | | | | | 1000 | 66 | 22 | 25 | 25 |
| | | 0.4 | 24 | 2.08 | 0.21 | 1.28 | 0.08 | 0.0052 | 6.07E-06 | 0.0104 | 2.03E-05 | 0.0065 | 8.85E-06 | 100 | 88 | 38 | 30.16 | 30.16 |
| | | | | | | | | | | | | | | 1000 | 17 | 5 | 19.05 | 22.73 |
| | 0.4 | 0.2 | 30 | 1.93 | 0.32 | 1.55 | 0.19 | 0.0051 | 8.26E-06 | 0.0092 | 2.00E-05 | 0.0075 | 1.42E-05 | 100 | 439 | 187 | 33.28 | 29.87 |
| | | | | | | | | | | | | | | 1000 | 86 | 17 | 19.63 | 16.5 |
| | | 0.4 | 30 | 1.93 | 0.36 | 1.23 | 0.15 | 0.0052 | 8.51E-06 | 0.0094 | 2.05E-05 | 0.0061 | 9.68E-06 | 100 | 110 | 49 | 32.1 | 30.82 |
| | | | | | | | | | | | | | | 1000 | 22 | 5 | 15.38 | 18.52 |
| | 0.1 | 0.2 | 56 | 1.28 | 0.61 | 1.05 | 0.24 | 0.0044 | 5.28E-06 | 0.0047 | 4.08E-06 | 0.004 | 3.13E-06 | 100 | $\geq$ 1000 | - | - | - |
| | | | | | | | | | | | | | | 1000 | 189 | 62 | 33.22 | 24.7 |
| | | 0.4 | 55 | 1.3 | 0.52 | 0.89 | 0.2 | 0.0044 | 5.54E-06 | 0.0048 | 4.38E-06 | 0.0034 | 2.48E-06 | 100 | 314 | 219 | 44.72 | 41.09 |
| | | | | | | | | | | | | | | 1000 | 48 | 13 | 27.27 | 21.31 |

Table S17: **Full Simulation Results for $n = 10000$, One-Sample t-test With Matching On Sampled Non-Infectious Individuals.** $R_0$ = target $R_0$. $k$ = overdispersion. $\Delta$ = % reduction in transmission. $t$ = day of intervention. $R_t^0 = R_t$ without treatment. $R_t^1 = R_t$ with treatment. $I_t$ = proportion of infectious persons at day of intervention. $I_{t+1}^0$ = proportion of infectious persons at time of sampling without treatment. $I_{t+1}^1$ = proportion of infectious persons at time of sampling with treatment. $m$ = number of persons sampled from each cluster. $C_S$ = number of clusters needed for each arm from simulations.$C_S^0$ = number of clusters needed for each arm from simulations using a two-sample Welch's t-test in Table S3. $C_S^M$ = number of clusters needed for each arm from simulations using a two-sample Weclh's t-test in Table S14.
